# lncSigAML: An AI-derived Long Non-coding RNA Prognostic Signature with Multi-Cohort Validation across 2,681 with Acute Myeloid Leukemia Patients

**DOI:** 10.64898/2026.11.17.26367793

**Authors:** Lan Thi-Ngoc Trinh, Thuy-An Nguyen, Minh-Khang Le, Tan-Hoang Nguyen, Stefan Bernadiskov, Danh-Tai Hoang, Bon Q. Trinh

## Abstract

Acute myeloid leukemia (AML) is clinically and genetically heterogeneous. Current risk stratification systems based on cytogenetic and genetic abnormalities do not fully capture prognostic heterogeneity, particularly within intermediate-risk groups. Long non-coding RNAs (lncRNAs) have emerged as key regulators of AML biology, contributing to leukemogenesis, impaired differentiation, apoptosis resistance, and treatment response. In this study, we developed lncSigAML, a bone marrow-derived lncRNA prognostic signature, using in pediatric samples from TARGET-AML (n = 1,594) and validated it in four bone marrow and peripheral blood datasets from TARGET-AML, Beat AML, and TCGA-LAML (n = 1,087). Ten lncRNAs were selected, including *MEF2C-AS1*, *LINC00477*, *PROSER2-AS1*, *LINC01122*, *KCNJ2-AS1*, *LINC01108*, *AP001042.1*, *SOCS2-AS1*, *ST3GAL6-AS1*, and *LINC00355*. Among candidate models, random survival forest achieved the best performance and significantly stratified patients into high- and low-risk groups (cross-validated HR = 2.87, 95%CI: 2.43-3.4; *P* < 0.001) in TARGET-AML bone marrow samples, with stable prognostic performance over 1-, 3-, and 5- year follow-up. The lncSigAML signature also robustly demonstrates significant prognostic effects across all cross-cohort and cross-tissue datasets separately (pooled HR = 2.91; 95%CI = 2.31-3.68; *P* <0.001). Meta-analysis showed substantially higher pooled 1-, 3-, and 5-year overall survival in the low-risk group than in the high-risk group (80.5%, 63.4%, and 55.3% vs. 52.1%, 28.1%, and 23.9%, respectively). Multivariable Cox regression demonstrated that lncSigAML provided prognostic information beyond clinical variables, genetic subtypes, and cytogenetic risk, with significant incremental prognostic value by partial likelihood ratio testing. Pathway analysis showed enrichment of genes related to cell survival and proliferation, impaired immune surveillance, and aggressive AML biology in the high-risk group, including STK33-, IL2, and PTEN-associated pathways. To support clinical and research applications, we implemented lncSigAML as an interactive, publicly accessible web application for individualized prognostic estimation. Prospective validation and functional studies of the identified lncRNAs will further evaluate their potential utility in precision AML care.

**KEY POINTS:**

- A 10-lncRNAsignature derived from 1,594 pediatric AML bone marrow samples stratified survival across 4 validation datasets.
- A lncRNA-based survival signature was constructed and deployed to a web application.

## INTRODUCTION

Acute myeloid leukemia (AML) is a biologically heterogeneous hematologic malignancy characterized by abnormal proliferation and impaired differentiation of myeloid blasts, diverse genetic alterations, variable treatment responses, and markedly different clinical outcomes. Current risk stratification systems, including the European LeukemiaNet (ELN), recommend incorporating recurrent cytogenetic and genetic abnormalities to define prognostic risk groups ^1,2^. Although these molecular classifications have substantially improved AML management, they still do not fully capture prognostic heterogeneity, particularly among intermediate-risk patients ^2,3^. This has motivated the search for complementary biomarkers that can capture prognostic information beyond established clinical and genetic risk factors.

Long non-coding RNAs (lncRNAs) are involved in biological processes that are important for tumorigenesis and cancer progression, including cell cycle regulation, proliferation, apoptosis, migration, and genome stability ^4,5^. They can regulate gene expression through multiple mechanisms, including chromatin remodeling, transcription, RNA splicing, RNA stability, and translation ^6,7^. In hematopoiesis and leukemia, lncRNAs have been linked to proliferation, differentiation, therapy resistance, and treatment outcomes ^8,9^. These functions suggest that lncRNA expression may capture biological features of AML that are not fully reflected by genetic alterations. Therefore, studying lncRNAs may provide additional prognostic information for patients with AML.

Many studies have identified individual lncRNAs and lncRNA-based signatures associated with survival in AML (Li, Li et al. 2022, Xie, Chen et al. 2023, Zhu, He et al. 2023, Seifpour, Zomorrod et al.2025, Pavlovic, Tosic et al. 2026). However, limitations remain in the development and validation of lncRNA-based prognostic models for AML. Several published signatures were developed from a single cohort, relatively small patient populations, or the TCGA-LAML dataset (Liu, Guo et al. 2021, Li, Li et al.2022, Li, Wu et al. 2022, Qin, Li et al. 2024). In addition, some studies have limited or no external validation, making it difficult to determine whether their prognostic findings can be reproduced in independent patient cohorts (Li, Li et al. 2022, Zhu, He et al. 2023). Many models also did not adjust for important clinical, genetic, or cytogenetic risk factors, and their prognostic value may therefore overlap with established risk factors.

In this study, we developed and validated lncSigAML, a bone marrow-derived lncRNA signature for AML prognosis. We used TARGET-AML bone marrow samples as the discovery cohort and applied a multistep approach to identify prognostic lncRNAs. We benchmarked six survival prediction methods using cross-validation and selected the best-performing model to construct lncSigAML. We then tested lncSigAML in four independent bone marrow and peripheral blood datasets from TARGET-AML, Beat AML, and TCGA-LAML. We also assessed its calibration, performance across validation cohorts, robustness to incomplete lncRNA measurements, and prognostic value after adjustment for clinical, genetic, and cytogenetic risk factors. Finally, we investigated the biological pathways associated with distinct lncSigAML risk groups and developed a publicly accessible web application for estimating lncSigAML risk from RNA-sequencing data.

## METHODS

### Study cohorts

Detailed information on sample collection is provided in the Supplementary Method. This study included three independent AML cohorts: TARGET-AML, Beat AML, and TCGA-LAML. From these cohorts, we divided the patients into five non-overlapping datasets according to the tissue profiled by RNA-seq: bone marrow from TARGET-AML (BM1) and Beat AML (BM2), and peripheral blood from TARGET-AML (PB1), Beat AML (PB2), and TCGA-LAML (PB3). In TARGET-AML (BM1), 1,594 AML bone marrow samples from 1,594 patients were used for lncRNA discovery and development of lncSigAML, while an independent set of 356 matched bone marrow samples from 356 Beat AML patients (BM2) was used for same-tissue external validation. Peripheral blood samples were used for cross-tissue validation of lncSigAML. After data preprocessing and sample matching, peripheral blood samples were available from 312 primary AML patients in TARGET-AML (PB1), 277 patients in Beat AML (PB2), and 142 patients in TCGA-LAML (PB3). Within each cohort (TARGET-AML and Beat AML), the patients included in the matched bone marrow (BM1/BM2) and peripheral blood (PB1/PB2) datasets were mutually exclusive; that is, no patient contributed samples to both the bone marrow and peripheral blood analyses.

### RNA-seq preprocessing and lncRNA annotation

Expression matrices were organized with genes as rows and samples as columns and were restricted to samples with matched clinical annotations. The Beat AML (BM2 and PB2) full expression matrix contained 63,677 genes, while the TARGET-AML and TCGA-LAML (BM1, PB1, and PB3) full expression matrices contained 60,660 genes. Differences in gene identifiers and the number of genes among the Beat AML, TARGET-AML, and TCGA-LAML datasets arise from the use of different reference genome assemblies and gene annotation versions. Beat AML used GRCh37 (hg19), whereas TARGET-AML and TCGA-LAML used GRCh38 (hg38). For Beat AML, duplicate gene symbols were made unique before downstream filtering. lncRNAs were identified using dataset-provided gene biotype annotations. Genes annotated as lincRNA, antisense, processed transcript, sense intronic, sense overlapping, or 3’ overlapping ncRNA were retained as lncRNA-related transcripts, resulting in 14,643 lncRNA genes. For TARGET-AML and TCGA-LAML, genes annotated with the lncRNA biotype were retained, resulting in 16,901 lncRNA genes. Because gene annotation schemes differed between datasets, lncRNA filtering was performed using cohort-specific biotype labels provided with each expression dataset ^10^. Details on methodologies are provided in Supplemental Materials.

## RESULTS

### Study Overview

We developed lncSigAML, a machine learning-based lncRNA signature, in TARGET-AML bone marrow RNA-seq data (BM1) and validated it on four independent datasets from three cohorts, including TARGET-AML (PB1), Beat AML (BM2 and PB2), and TCGA-LAML (PB3) (Fig. 1A). We first performed multi-step prognostic screening in BM1 and identified an optimal set of 10-prognostic lncRNA signature for AML (lncSigAML), including *MEF2C-AS1*, *LINC00477*, *PROSER2-AS1*, *LINC01122*, *KCNJ2-AS1*, *LINC01108*, *AP001042.1*, *SOCS2-AS1*, *ST3GAL6-AS1*, and *LINC00355* (Fig. 1B). This process was followed by 5-fold cross-validation to select the optimal ML approach, with random survival forest (RSF) achieving the best performance (Fig. 1C-D). The resulting lncSigAML model was then evaluated across four independent datasets spanning bone marrow and peripheral blood samples, where it consistently demonstrated prognostic significance and provided additional prognostic value beyond established clinical, genetic and cytogenetic risk factors, as demonstrated by multivariable Cox regression and partial likelihood ratio tests (Figs. 1E-F).

**Figure 1.**
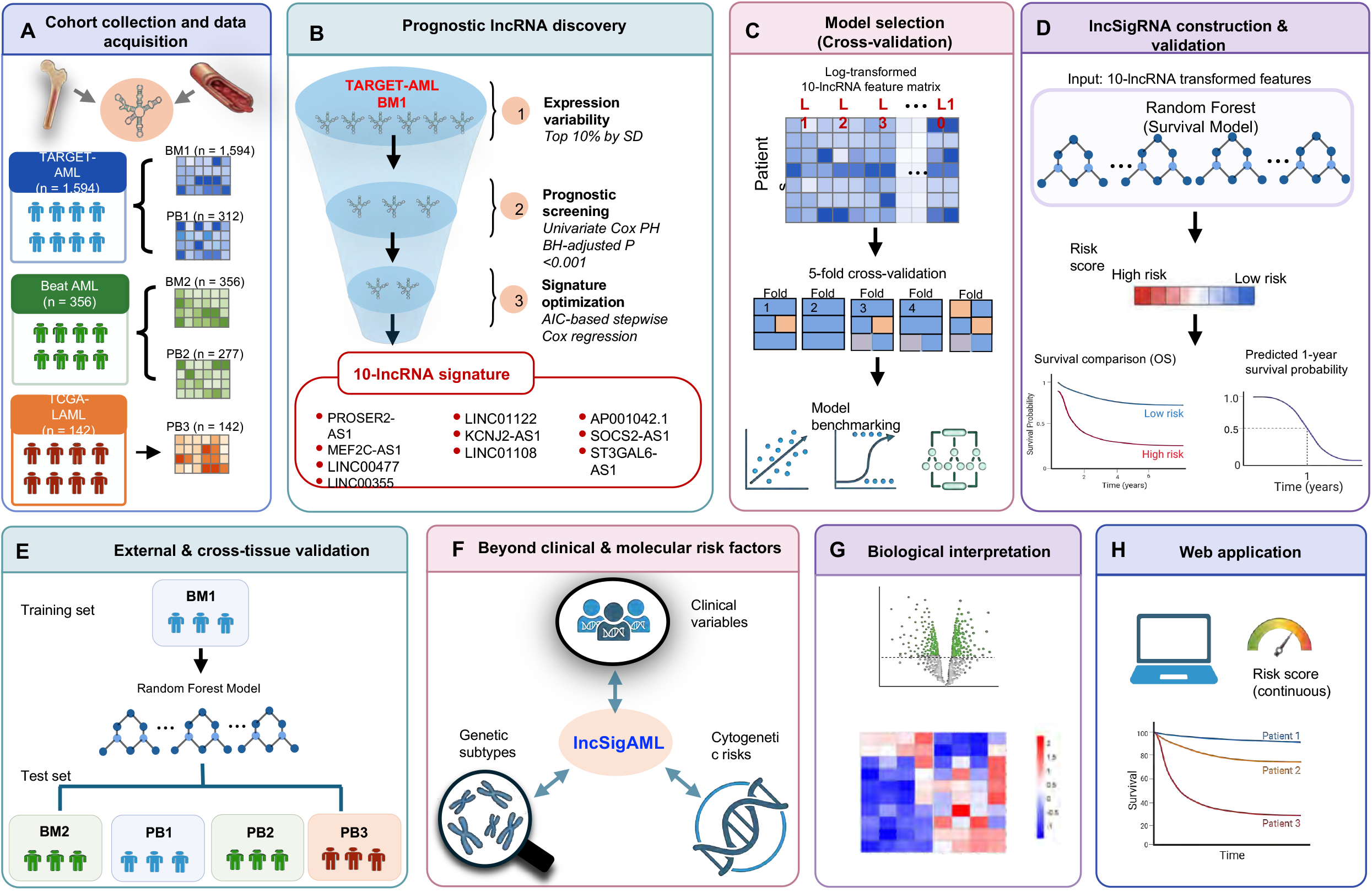
Overall design of the study. (A) Three independent AML cohorts comprising five datasets derived from bone marrow and peripheral blood samples. (B) Multi-step feature selection to identify the 10-lncRNA lncSigAML signature in the BM1 cohort. (C) Benchmarking of six survival prediction models using five-fold cross-validation to identify the best-performing model for lncSigAML. (D) Evaluation of the selected random survival forest model in the BM1 cohort, including risk stratification, survival analysis, calibration, and time-dependent receiver operating characteristic analysis. (E) External validation of lncSigAML using a model trained on the BM1 cohort and independently tested across BM2, PB1, PB2, and PB3 cohorts. (F) Evaluation of the prognostic value of lncSigAML beyond clinical features, genetic alterations, and cytogenetic risk. (G) Biological characterization of lncSigAML-associated risk groups using differential expression, heatmap, and C6 pathway enrichment analyses. (H) Overview of the lncSigAML web application.

Pathway analyses identified significant enrichment of gene sets related to oncogenic signaling (KRAS/STK33), immune and hematopoietic signaling (IL-2/IL-21/LEF1), proliferation and cell-state regulation (MEL18/RB-p130), and metabolic/translational regulation (mTOR/EIF4GI/BCAT) in AML bone marrow samples associated with lncSigAML. These findings suggest that lncSigAML captures a distinct AML transcriptional state characterized by coordinated remodeling of oncogenic, immune, proliferative, and metabolic programs (Fig. 1G). To facilitate the broad accessibility and potential clinical application of lncSigAML, we developed a user-friendly interactive web application that enables users to calculate lncSigAML scores from either individual patient-level lncRNA expression profiles or batch expression data from multiple patients in CSV format, with scoring results available for download (Fig. 1H).

### Multi-step screening of 1,594 bone marrow transcriptomes identifies a 10-prognostic signature (lncSigAML)

Because AML originates in the bone marrow, we began by conducting a multistep prognostic screen of bone marrow transcriptomes to identify top prognostic lncRNAs. Using the comprehensive TARGET-AML dataset (dbGaP Study Accession: phs000465.v23.p8), we compiled the BM1 cohort of bone marrow samples from 1,594 patients (Table S1 and Fig. 1A). By applying unsupervised filtering based on the standard deviation of log-transformed read counts, we identified 217 highly variable lncRNAs across samples (Table S2). We next performed supervised screening using univariate Cox proportional hazards analyses on these lncRNAs with multiple hypothesis correction (Bonferroni) and identified the 24 most significant lncRNAs associated with overall survival (OS-associated lncRNAs) (adjusted *p* < 0.001) (Fig. S1A). We then applied a stepwise saturation algorithm based on the Akaike Information Criterion (AIC) with Cox regression model to the 24 OS-associated lncRNAs (Fig. S1A), and identified an optimal set of 10 lncRNAs, beyond which additional lncRNAs provided minimal improvement in model fit and increased the risk of overfitting (Figs. 1B and 2A). Among the identified lncRNAs, six were antisense to nearby protein-coding genes; five overlapped the corresponding gene body and promoter region, whereas one was divergently transcribed (Figs. 2A and S1B-H). Four were intergenic lncRNAs, with two located upstream and two downstream of their respective neighboring coding genes (Figs. 2A and S1I-L). Of note, high expression of eight lncRNAs was associated with poor overall survival (OS), whereas low expression of two lncRNAs was associated with poor OS (Figs. 2A and S1B). We hereafter refer to this set of OS-associated lncRNAs as the lncRNA prognostic signature for AML (lncSigAML).

**Figure 2.**
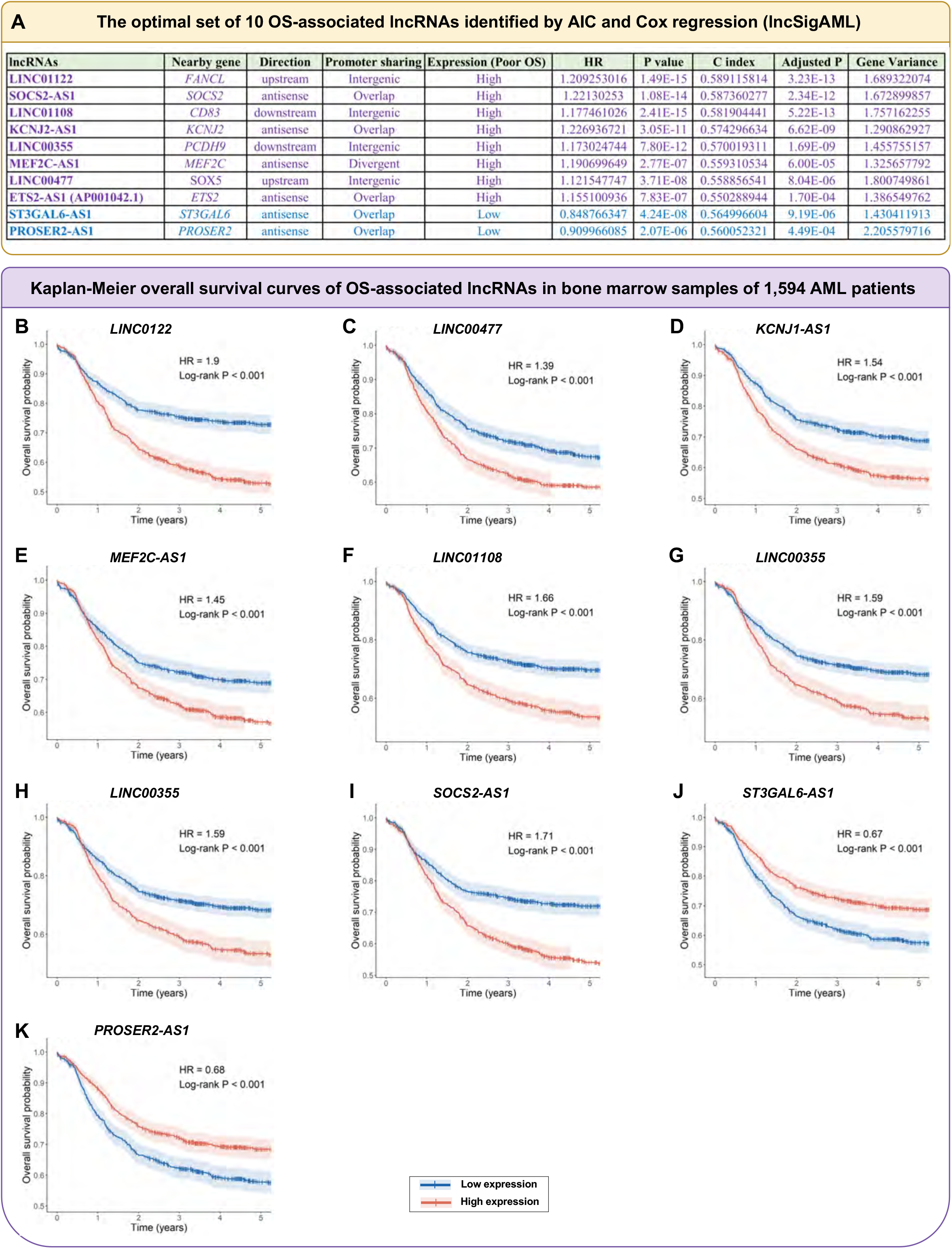
Identification of 10-OS-associated lncRNAs signature lncSigAML. (A) Characteristics of the 10 lncRNAs selected from the 24 OS-associated lncRNA candidates using stepwise Cox regression with AIC-based model selection. The table summarizes the genomic relationship of each lncRNA with its nearby gene, expression direction associated with poor OS, hazard ratio (HR), P value, concordance index (C-index), Bonferroni-adjusted P value, and expression variance across BM1 samples. (B-K) Kaplan-Meier analyses comparing overall survival between low- and high-expression groups for each selected lncRNA. See also Figure S1 and Table S3.

### Construction of the lncSigAML-based machine-learning model with a robust survival prediction

We evaluated the standardized lncSigAML feature matrix in the BM1 cohort using six commonly used machine-learning algorithms for survival prediction (Fig. 1C). These include Cox proportional hazards (CoxPH) regression ^11^, LASSO regression ^12^, Elastic Net regression ^13^, random survival forest (RSF) ^14^, XGBoost survival model ^15^, and GradientBoost ^16^. We performed five-fold cross-validation and calculated the mean and standard deviation of the Harrell’s concordance index (C-index) ^10^ across the five folds for each model (Fig. 1C). Among the six models evaluated, random survival forest demonstrated the highest predictive performance for computing lncSigAML signature, achieving the highest cross-validated C-index across the 5-fold cross-validation (mean C-index of 0.653 ± 0.023). This was followed by the standard CoxPH model (mean C-index = 0.649 ± 0.015), Elastic Net (mean C-index = 0.648 ± 0.015), and LASSO regression model (mean C-index = 0.648 ± 0.016), all of which exhibited comparable performance (Figs. 1C and 3A). GradientBoost and XGBoost yielded lower mean C-indices of 0.629 (± 0.020) and 0.612 (± 0.021), respectively (Fig. 3A).

**Figure 3.**
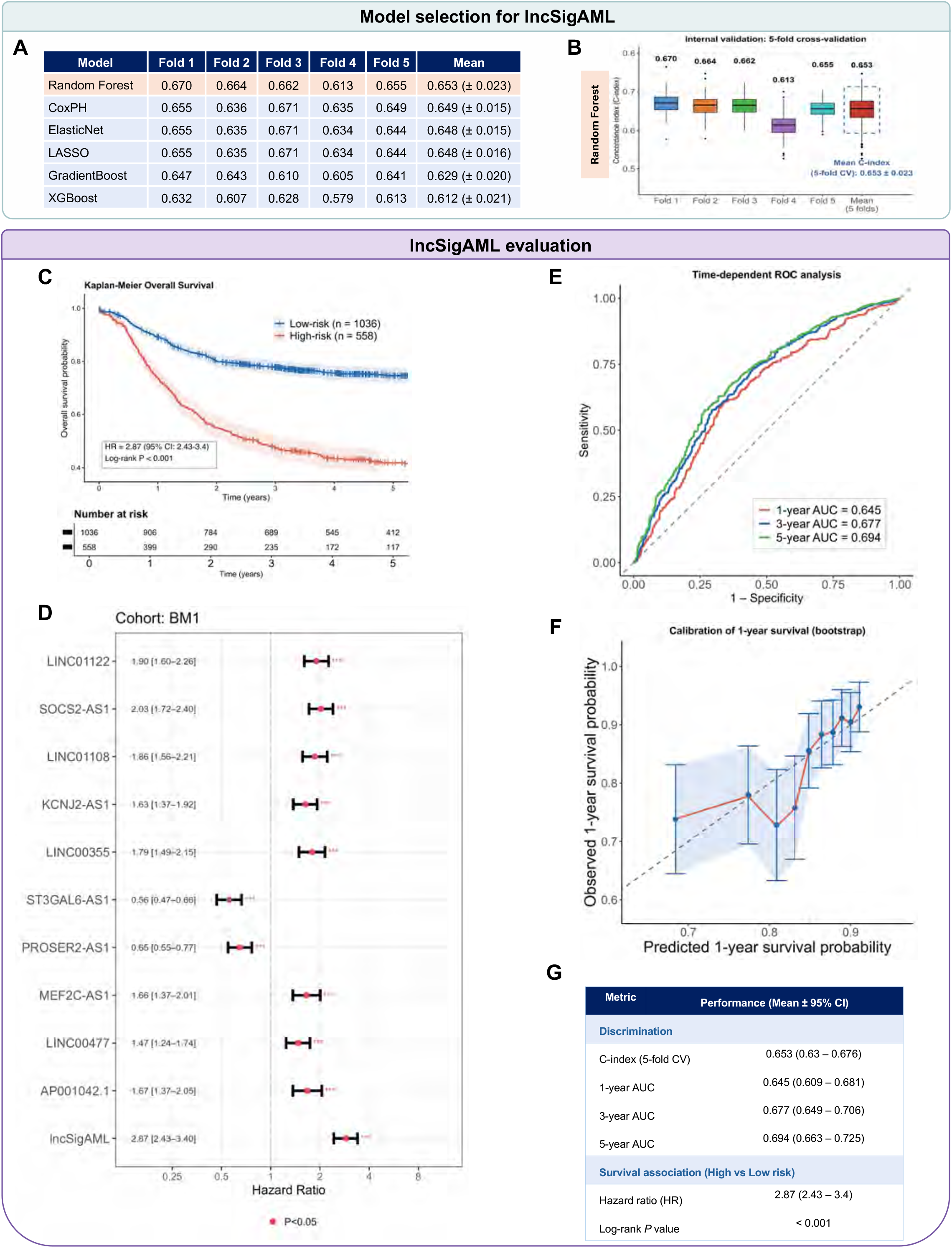
Model selection and evaluation of lncSigAML in the BM1 cohort. (A) Comparison of model performance during five-fold cross-validation in the BM1 cohort. C-index values are reported for each fold across six candidate models: Random Forest, CoxPH, Elastic Net, LASSO, Gradient Boosting, and XGBoost. The mean C-index across the five folds is shown in the final column, with values in parentheses representing the SD across folds. (B) Random Forest performance during five-fold cross-validation in the BM1 cohort. Boxplots show the distribution of C-index values for each fold, with numbers above the boxes indicating the mean C-index. The final boxplot summarizes performance across all five folds, with an overall mean C-index of 0.653 ± 0.023 (mean ± SD). (C) Kaplan-Meier analysis of overall survival in the BM1 cohort stratified into low- and high-risk groups based on the Random Forest-derived risk score. The low-risk group included 1,036 patients and the high-risk group included 558 patients. Shaded areas indicate 95% confidence intervals. The high-risk group showed significantly shorter overall survival than the low-risk group (HR, 2.87; 95% CI, 2.43-3.40; log-rank *P* < 0.001). (D) Forest plot showing the hazard ratios (HRs) and 95% confidence intervals for lncSigAML and its component lncRNAs in the BM1 cohort. The dashed vertical line indicates HR = 1; red points indicate *P* < 0.05; three asterisks indicate *P* < 0.001. (E) Time-dependent receiver operating characteristic (ROC) curves of the lncSigAML risk score for predicting 1-, 3-, and 5-year overall survival in the BM1 cohort. The corresponding areas under the curve (AUCs) were 0.645, 0.677, and 0.694, respectively. The dashed diagonal line indicates the reference line for random prediction. (F) Calibration plot for 1-year overall survival in the BM1 cohort. Predicted survival probabilities are compared with observed survival probabilities across risk groups. The solid diagonal line represents perfect calibration, and error bars indicate the 95% confidence intervals of the observed estimates. (G) Summary of lncSigAML performance in the BM1 cohort. Discrimination was assessed by the 5-fold cross-validated C-index and time-dependent AUCs at 1, 3, and 5 years. Survival association was evaluated by the hazard ratio (HR) comparing high- and low-risk groups and the log-rank test. Values are reported as mean with 95% confidence intervals (CI), except the log-rank *P* value. See also Figure S2 and Table S3.

To assess internal validity and generalizability, we performed 5-fold cross-validation of the <u>lncSigAML</u> model. The model achieved a mean C-index of 0.653 ± 0.023 (95% CI: 0.630–0.676), with fold-specific values ranging from 0.613 to 0.670, indicating stable discriminative performance across folds and suggesting that results were not driven by any single data subset (Fig. 3B). Next, to demonstrate the prognostic effect of the lncSigAML model, we stratified 1,594 AML patients (BM1 cohort) into low-risk (n = 1,036) and high-risk (n = 558) groups based on the maximally selected rank statistics (MSRS) that provides the strongest separation in overall survival between the resulting risk groups. Kaplan-Meier survival analysis confirmed a statistically significant difference in OS between the two risk groups (log-rank *P* < 0.001) (Fig. 3C), wherein high-risk patients exhibited markedly worse outcomes compared with their low-risk counterparts, corresponding to a hazard ratio of 2.87 (95% CI: 2.43–3.40; *P* < 0.001) (Fig. 3D). The 1-, 3-, and 5-year survival probabilities were 73.6% (70.0% - 77.4%), 47.5% (43.4% - 51.9%), and 41.9% (37.8%-46.4%) in the high-risk group versus 89.2% (87.4 - 91.1%), 77.8% (75.3% - 80.4%), and 74.6% (71.8% - 77.4%) in the low-risk group, respectively. This indicates that the high-risk lncSigAML group has substantially worse overall survival than the low-risk group, and the difference becomes particularly pronounced by 3–5 years.

We next assessed calibration, the agreement between predicted and observed 1-, 3-, 5-year survival. 1-, 3-, and 5-year calibration plots showed close agreement between predicted and observed survival probabilities across the full risk range. The bootstrap-corrected curve closely followed the ideal 45° line, indicating that lncSigAML provided well-calibrated survival predictions (Figs 3F and S2). Furthermore, time-dependent receiver operating characteristics (ROC) analyses at 1-year, 3-year, and 5-year time points showed AUCs of 0.645 (95%CI = 0.609 - 0.681), 0.677 (95%CI = 0.649 - 0.706), and 0.694 (95%CI = 0.663 - 0.725), respectively (Fig. 3G). The stability of AUC estimates across time points suggests that the prognostic signal encoded by lncSigAML is not confined to short-term risk prediction but persists over longer follow-up durations, underscoring the potential clinical utility of this model as a temporally consistent prognostic tool for patients with AML. Thus, we selectedrandom survival forest as the optimal model for subsequent analyses constructing the lncRNA-based prognostic model for AML patients.

### lncSigAML generalizes across cohorts, tissues, and platforms and tolerates missing lncRNA measurements

We next examined lncSigAML prognostic performance generated from our training set (BM1) across four test sets (Fig. 1E). These include the PB1 set of 312 peripheral blood (PB) samples from the TARGET-AML cohort, the BM2 set of 356 BM samples and the PB2 set of 277 PB samples from the Beat AML cohort, and the PB3 set of 142 PB samples from the TCGA-LAML cohort (Fig. 1A). In the external bone marrow (BM) validation set (BM2), Kaplan–Meier analysis demonstrated significantly different overall survival between the low- and high-risk groups (log-rank *P* < 0.001) (Fig. 4A). In line with this finding, we observed significantly different overall survival between the low- and high-risk groups (log-rank *P* < 0.001) in the internal peripheral blood (PB) set (PB1; Fig. 4B) and the external peripheral blood validation sets (PB2 and PB3; Figs. 4C-D). Consistently, high-risk patients exhibited significantly higher mortality than low-risk patients, with hazard ratios of 2.54 (95% CI, 1.60–4.03) in BM2 (Fig. 4E), 3.65 (95% CI, 2.39–5.57) in PB1 (Fig. 4F), 2.83 (95% CI, 1.60–5.01) in PB2 (Fig. 4G), and 2.71 (95% CI, 1.75–4.21) in PB3 (Fig. 4H). Moreover, a pooled meta-analysis across the four independent sets confirmed the robust prognostic value of lncSigAML (pooled HR = 2.91; 95% CI, 2.31–3.68; *P* < 0.001) (Fig. 4I). To further evaluate the generalizability of the lncSigAML across different sequencing platforms, we reversely constructed lncSigAML in the Beat AML BM set (BM2) and evaluated its prognostication in the TARGET-AML BM set (BM1). The model retained appreciable prognostic discrimination in this adult AML cohort, yielding a C-index of 0.637 (95% CI: 0.61–0.66) and a 1-year AUC of 0.644 (95% CI: 0.61–0.68) (Fig. 4J). In line with this finding, cross-platform lncSigAML calibration for predicted 1-, 3-, and 5-year overall survival probabilities in this independent BM2 dataset showed agreement between predicted and observed survival probabilities (Figs. S3A-C). Thus, lncSigAML preserved its prognostic performance across independent cohorts despite differences in sequencing platforms and tissue sources.

**Figure 4.**
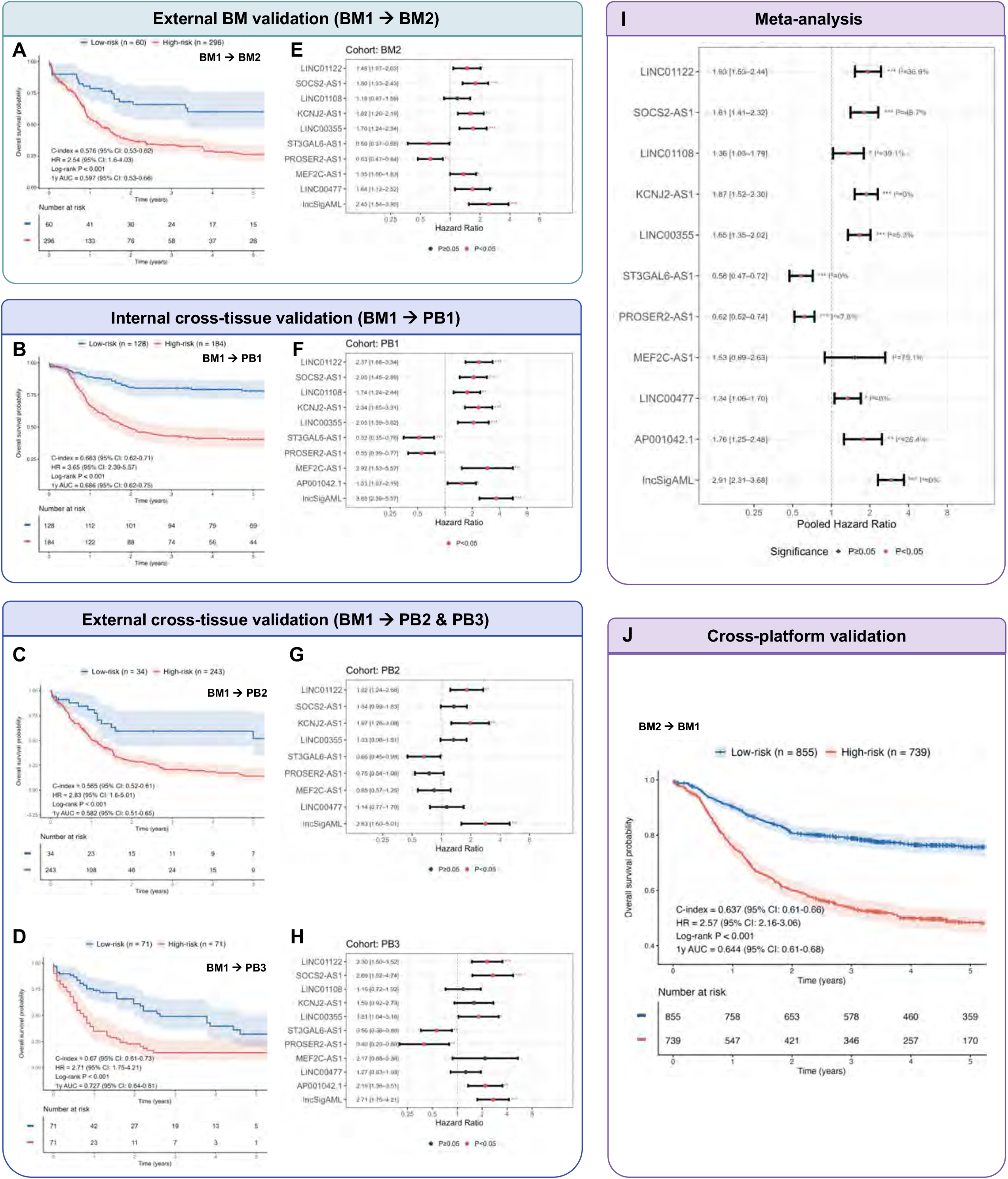
External, cross-tissue, and cross-platform validation of lncSigAML across independent AML cohorts. (A) Kaplan–Meier survival analysis for external validation of lncSigAML in the BM2 cohort. Patients were stratified into low- and high-risk groups using maximally selected rank statistics. Numbers at risk indicate patients remaining under observation at each time point. (B-D) Kaplan–Meier survival analyses for cross-tissue validation of lncSigAML in the PB1 (B), PB2 (C), and PB3 (D) datasets. Patients were stratified into low- and high-risk groups using maximally selected rank statistics. Numbers at risk indicate patients remaining under observation at each time point. (E - H) Hazard ratio forest plot for lncSigAML and its component lncRNAs in the BM2 (E), PB1 (F), PB2 (G), and PB3 (H). The dashed vertical line indicates HR = 1. Red points indicate statistically significant associations (*P* < 0.05), whereas dark points indicate *P* ≥ 0.05. One, two and three asterisks indicate *P* < 0.05, 0.01, and 0.001, respectively. (I) Pooled analysis of the prognostic effect of lncSigAML and its component lncRNAs across all independent validation cohorts (BM2, PB1, PB2, and PB3). The dashed vertical line indicates HR = 1. Red points indicate statistically significant associations (*P* < 0.05), whereas dark points indicate *P* ≥ 0.05. One, two and three asterisks indicate *P* < 0.05, 0.01, and 0.001, respectively. (J) Cross-platform validation of lncSigAML using the BM2 cohort to stratify patients in the BM1 cohort.Kaplan–Meier analysis demonstrated significantly different overall survival between the low- and high-risk groups (log-rank P < 0.001). Model performance is summarized by the concordance index (C-index) and 1-year area under the receiver operating characteristic curve (AUC), with 95% confidence intervals. Numbers at risk indicate patients remaining under observation at each time point. See also Figure S3 and Table S3.

To evaluate stability of lncSigAML to incomplete expression measurements, we performed missing-data sensitivity analysis (Fig. S3D). C-index and 1-year AUC estimates remained largely consistent as the proportion of missing data increased from 10% to 50%, with bootstrap-corrected 95% CI exhibiting substantial overlap across all tested thresholds (Figs. S3E-F). Additionally, calibration plots constructed under each missingness condition similarly indicated that bias-corrected bootstrap calibration curves maintained adequate agreement with the line of ideal calibration across increasing levels of data incompleteness at 1 (Figs. S3G-J), 3 (Figs. S3K-N), and 5 (Figs. S3O-R) years, respectively. Collectively, these sensitivity analyses indicate that imputation-based recovery of missing lncRNA expression values did not meaningfully distort probability estimates, and that the lncSigAML maintained stable discriminative performance across the full range of missingness conditions examined, supporting its robustness across different datasets and substantial levels of feature-level missingness.

### lncSigAML provides independent prognostic information beyond clinical and molecular risk factors

To further evaluate whether the lncRNA-derived risk score provided prognostic information independent of basic clinical characteristics, we performed multivariable Cox proportional hazards regression using cross-validated patient-level risk scores in the TARGET-AML bone marrow cohort (BM1). Specifically, each patient’s risk score was generated by the cross-validation version of lncSigAML random forest where the patient belongs to the held-out test fold, ensuring that the prediction for every individual was obtained from a model that had not been trained on that patient’s data. This approach minimized information leakage and provided a less biased estimate of the prognostic value of the lncRNA signature. The cross-validated lncRNA risk score remained independently associated with overall survival after adjustment for age, sex, and race (HR = 2.62, 95% CI: 2.20–3.13, *P* < 0.001) (Table 1). Multivariable analyses of other datasets (BM2, PB1-3) yielded consistent findings (Table S3). Thus, lncSigAML provides prognostic information beyond clinical variables.

Given that AML patients with different genetic subtypes (including cytogenetic and molecular classifications) exhibit distinct clinical outcomes and survival probabilities ^17^, we investigated whether the prognostic value of lncSigAML was confounded by underlying genetic subtype. The Beat AML bone marrow (BM2) cohort contains comprehensive genomic annotation, enabling adjustment for this important clinical variable (Table S1). We therefore performed a multivariable Cox proportional hazards analysis including both the lncSigAML risk group and AML genetic subtype as covariates. After adjusting for genetic subtypes, lncSigAML remained an independent predictor of overall survival. Patients classified into the high-risk group exhibited a significantly increased risk of mortality compared with those in the low-risk group (HR = 1.91, 95% CI: 1.14 - 3.21, *P* = 0.014) (Table 2). These findings demonstrate that the prognostic value of lncSigAML is not merely attributable to differences in the distribution of AML genetic subtypes but instead provides additional prognostic information beyond established genomic classification. A similar result was found in the Beat AML peripheral blood (PB2) cohort (Table S4). The partial likelihood ratio (PLR) test showed that lncSigAML adds a statistically significant prognostic value to the current genetic classification (*P* = 0.009, BM2; *P* = 0.019, PB2) (Table S4). These findings suggest that lncSigAML provides prognostic information beyond AML genetic subtype.

Because cytogenetic risk is a major prognostic factor ^17^, we further assessed whether the prognostic value of lncSigAML was independent of established cytogenetic risk. Since cytogenetic risk is available in PB3 dataset, we performed a multivariable Cox proportional hazards analysis including both the lncSigAML risk group and cytogenetic risk as covariates in the TCGA-LAML peripheral blood (PB3) cohort. Consistent with the findings in the Beat AML cohort, patients classified into the lncSigAML high-risk group remained at significantly greater risk of mortality after adjustment for cytogenetic risk (HR = 2.12, 95% CI: 1.307–3.453, P = 0.002). PLR test showed that lncSigAML adds a statistically significant prognostic value to the current cytogenetic risk category (*P* = 0.002, PB3) (Table 3). These findings indicate that lncSigAML provides prognostic information beyond conventional AML cytogenetic risk classification.

### High-risk lncSigAML is associated with enhanced proliferative capacity, increased immune and microenvironmental signaling, and metabolic remodeling in AML

To assess the biological role of and provide scientific basis for future experimental examination of lncSigAML, we first examined expression pattern of lncSigAML lncRNA members in low-risk and high-risk AML groups across test datasets. Consistence with our finding of high expression associated with poor survival (Fig. 2), high expression of eight lncRNAs was associated with poor overall survival (OS), expression of these genes is up-regulated in high-risk groups across the data sets (Fig. 5A). Similarly, low expression of two lncRNAs was associated with poor OS (Fig. 2) and these genes are downregulated in high-risk groups across the data sets (Fig. 5A). We next performed differential expression (DEA) analysis between low- and high-risk lncSigAML groups and identified a repertoire of genes differentially expressed between risk groups (Fig. 5B, Table S5). Meta-analysis of gene-level differential expression ^18,19^ results identified the top 50 genes with consistent differential expression across independent test cohorts showing large expression differences (all adjusted *P* < 0.001) (Fig. 5C, Table S5). These genes included hematopoietic and transcriptional regulators (*MECOM*, *HLF*, *HOXA11*, *HOXA13*), immune and cytokine signaling genes (*SOCS2*, *IL12RB2*, *IL12A-AS1*, *KLRF2*, *NCR2*), and genes involved in developmental and growth factor signaling (*GLI2*, *FGF10*, *GPC4*, *SPOCK3*) (Fig. 5C).

**Figure 5.**
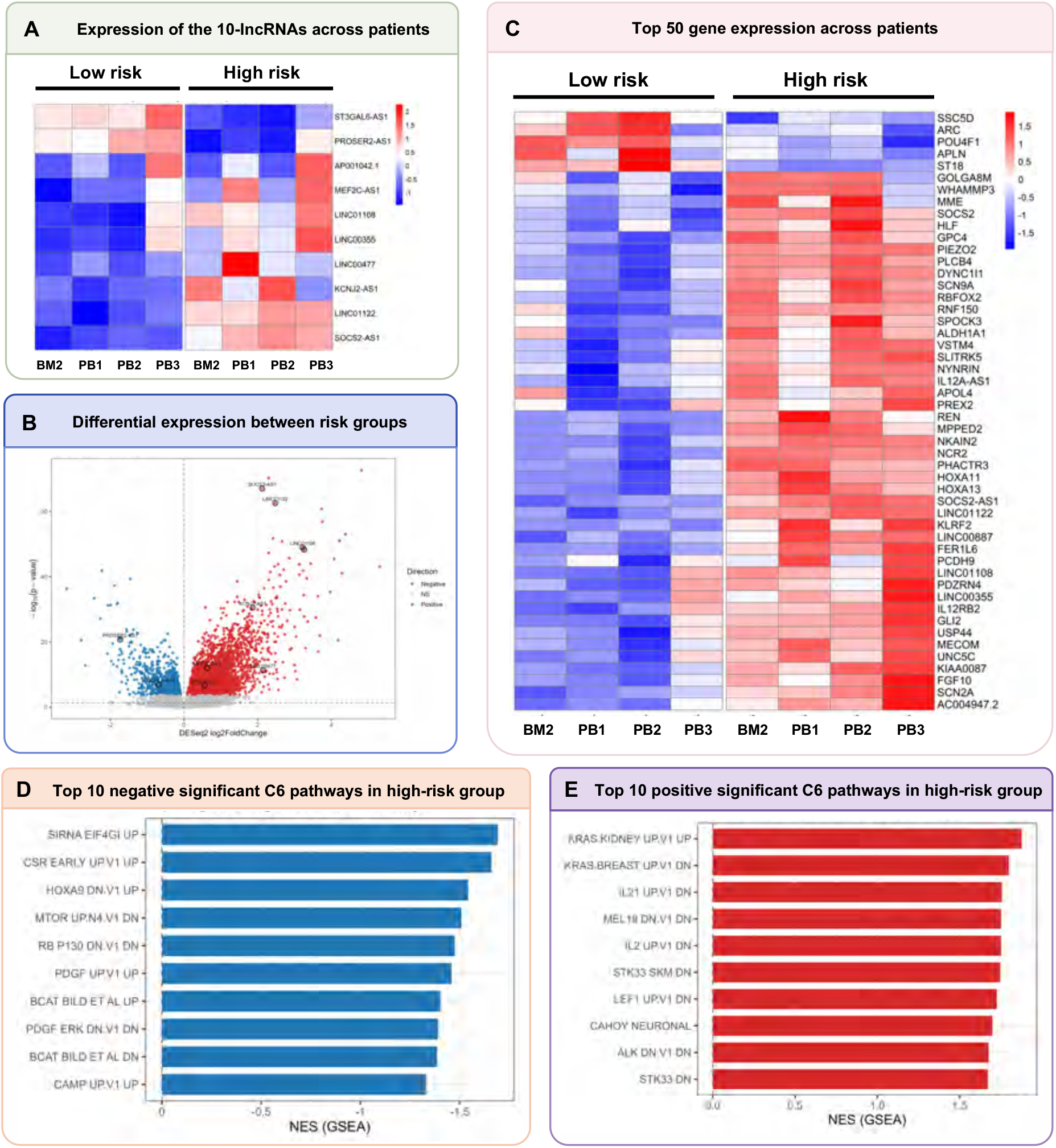
Gene expression and pathway analyses. (A) Heatmap demonstrating mean expression of the selected lncRNAs in low- and high-risk groups of the test cohorts using DESeq2 across test datasets. (B) Volcano plot showing the meta-analysis results of cohort-specific differential expression analyses using DESeq2. (C) Heatmap demonstrating the 50 top differentially expressed genes (DEGs) in low- and high-risk groups of the test cohorts. (D-E) Bar plots describing the top negative (D) and positive (E) C6 pathways in the high-risk group. See also Table S3.

To investigate the oncogenic processes associated with the lncSigAML risk signature, we performed gene set enrichment analysis (GSEA) ^20^ comparing the high-risk and low-risk AML groups using the C6 category of the MSigDB database (https://www.gsea-msigdb.org). The top negatively enriched pathways included translational and growth-regulatory processes, including EIF4GI, mTOR, RB/p130, and PDGF-associated signatures (NES = −1.46 to −1.69; adjusted P < 0.01) (Fig. 5D, Table S6). On the other hand, the high-risk group showed significant enrichment of pathways involved in cellular proliferation, immune signaling, and ion channel activity (Fig. 5E, Table S6). In particular, there was significant enrichment of transcriptional programs related to oncogenic signaling (KRAS/STK33; NES = 1.67–1.88), immune and hematopoietic signaling (IL-2/IL-21/LEF1; NES = 1.73–1.76), and proliferation and cell-state regulation (MEL18; NES = 1.75), with the leading positively enriched programs showing adjusted P values ranging from 2.9 × 10⁻⁷ to 1.7 × 10⁻⁵ in lncSigAML high-risk group (Fig. 5E, Table S6). Collectively, these findings suggest that lncSigAML captures an AML transcriptional state associated with increased cellular proliferation, cytokine signaling, and growth-factor activity, along with broader oncogenic and regulatory alterations.

### An interactive web application for personalized prognostic assessment

To facilitate clinical translation and broaden accessibility of lncSigAML as an RNA-based machine learning prognostic model, we developed lncSigAML webapp, an interactive web-based application using the R Shiny framework (Fig. 6). lncSigAML webapp enables users to obtain individualized prognostic assessments directly from patient-level RNA expression data without requiring specialized bioinformatics expertise or local software installation. The web interface comprises four sections: Overview, Application, Survival Table, and Instructions (Fig. 6A). The Overview provides a concise introduction to the lncSigAML framework, which predicts overall survival from lncRNA expression profiles. It summarizes the model architecture, including the Random Forest–based prediction approach, training cohort, input features, and cross-validation performance. A schematic of the study workflow and model development process is also provided. The Application provides an interactive interface for patient-specific survival prediction. Users can enter raw read-count expression values for the 10 lncRNAs comprising the lncSigAML signature and initiate prediction by clicking the Analyze Patient button. The application also supports uploading expression datasets in csv, txt, tsv, or excel format for batch analysis. Users can clear entered values and process multiple patient samples through the uploaded dataset. Users can also remove the currently entered patient data using the delete function. Following prediction, three analytical modules—Survival Curves, Risk Profile, and Expression Profile—provide complementary views of the predicted survival probability, risk classification, and expression patterns of the signature lncRNAs. The Survival Table provides the predicted survival probabilities in tabular form, while the Instructions section provides guidance on data input, file formatting, and use of the prediction functions (Fig. 6A). The clinical workflow begins with blood or bone marrow samples collected from patients, followed by RNA sequencing to generate raw read-count expression data. These data are uploaded to the lncSigAML webapp for individualized prognostic assessment (Fig. 6B). Within the application, the raw expression data are first filtered to extract the 10 lncRNAs comprising the lncSigAML signature and subsequently subjected to log1p transformation. The processed expression values are then entered into the pretrained Random Forest model embedded in the application backend to calculate an individualized lncSigAML risk score. Based on the resulting risk score, patients are classified into low-, intermediate-, or high-risk groups (Fig. 6C). The application dynamically generates individualized prognostic outputs, including the assigned risk level, predicted survival probabilities at key clinical time points (6 months, 1, 3, 5, and 10 years), predicted survival curves, and expression profiles of the 10 signature lncRNAs (Fig. 6D). Users can also enter multiple patients to compare predicted survival trajectories and review or export numerical survival estimates. Together, these features provide an accessible interface for translating the lncSigAML prognostic model into an individualized risk assessment framework.

**Figure 6.**
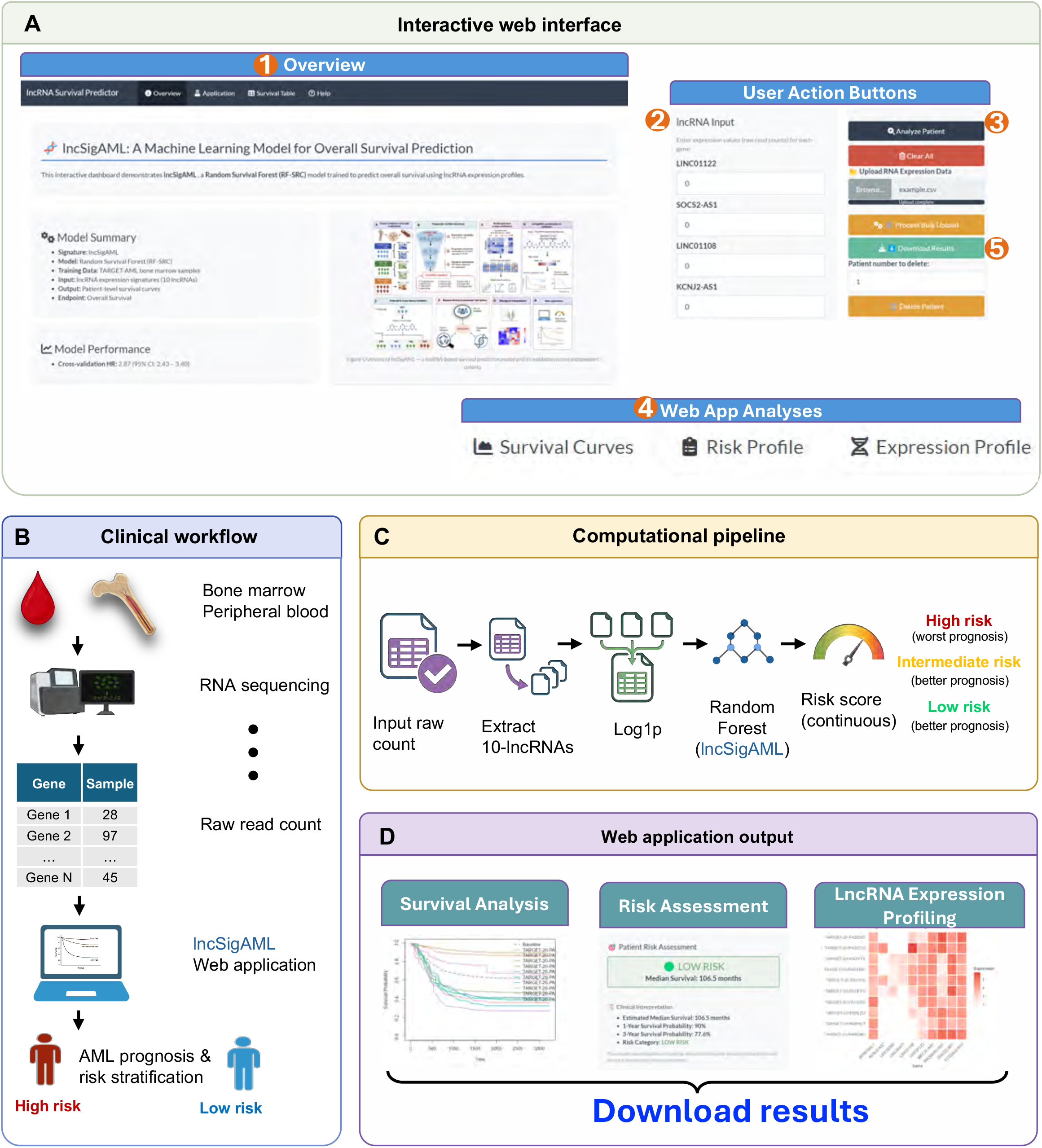
lncSigAML web application. (A) Web application interface. (1) Overview of the web interface. User action buttons include (2) entering lncRNA read-count values, (3) analyzing or deleting patient records, (4) viewing the prognostic results, (5) downloading the results. (B) Overall workflow of the lncSigAML web application. RNA sequencing is performed on bone marrow or peripheral blood samples from AML patients to generate raw read-count expression data, which are subsequently uploaded to the lncSigAML web application for individualized prognostic assessment. The resulting prognostic outputs can be reviewed and downloaded by users. (C) RNA expression processing and computation. Raw read-count expression data are extracted and subjected to log1p transformation and then input to the random survival forest model. The resulting risk score is used for downstream risk stratification and survival evaluation. (D) The output of the web application, including the predicted survival curve, risk assessment, survival probabilities, and lncRNA expression profiles.

In summary, lncSigAML provided robust prognostic stratification across independent AML cohorts and tissue sources, with prognostic information beyond established clinical, genetic, and cytogenetic risk factors. The high-risk lncSigAML state was associated with coordinated oncogenic, immune, proliferative, and metabolic programs, and was implemented in an accessible web application for individualized prognostic assessment (Fig. 1).

## DISCUSSION

In this study, we developed and validated a bone marrow-derived lncRNA prognostic framework for acute myeloid leukemia using transcriptomic profiles from five datasets and three independent cohorts comprising 2,681 pediatric and adult patients. By conducting a multi-step lncRNA prognostic screening, we established a ten-lncRNA signature that demonstrated consistent prognostic performance across independent datasets. The selected lncRNAs include *MEF2C-AS1*, *LINC00477*, *PROSER2-AS1*, *LINC01122*, *KCNJ2-AS1*, *LINC01108*, *AP001042.1*, *SOCS2-AS1*, *ST3GAL6-AS1*, and *LINC00355*. The final random survival forest model maintained stable prognostic discrimination through multiple external validation and sensitivity analyses, while remaining independently associated with overall survival after adjustment for basic clinical variables and genetic subtypes, including cytogenetic and molecular classifications. To facilitate reproducibility and accessibility, the final model was implemented as a publicly available web application. Collectively, these findings suggest that the signature derived from these selected lncRNAs was reproducibly prognostic.

lncRNAs have emerged as important regulators of hematopoiesis and leukemogenesis through their involvement in chromatin remodeling ^21^, transcriptional regulation ^22^, RNA stability ^23^, and signaling pathways ^24,25^. Consistent with previous reports, our findings further support the prognostic value of lncRNA expression in AML. Rather than relying on a single biomarker, we developed a signature derived from ten identified prognostic lncRNAs, many of which have established roles in AML biology. This combination of established and relatively unexplored lncRNAs suggests that clinically relevant prognostic information may arise from coordinated transcriptional programs rather than individual biomarkers alone. Further functional studies are warranted to elucidate the biological mechanisms through which these lncRNAs contribute to AML progression and treatment response.

Several lncRNAs within the signature, including *SOCS2-AS1* ^26^, *LINC01108* ^22^, and *AP001042.1* (known as LINC00265) ^27,28^ have previously been implicated in cancer biology or hematologic malignancies, whereas others remain poorly characterized. We found the opposite evidence of *LINC01108* as a biomarker of poorer prognosis across datasets compared to a previous study ^22^. While *MEF2C* plays a well-characterized, prominent oncogenic and prognostic role in acute leukemia ^29^, direct evidence specifically linking *MEF2C-AS1* to the pathogenesis of acute myeloid leukemia (AML) is sparse. Other lncRNAs, such as LINC00355 ^30^, *PROSER2-AS1* ^31^, *LINC01122* ^32^, and *ST3GAL6-AS1* ^33^ are also known to have functional roles in cancer although their explicit mechanisms in AML have not been explored.

An important strength of this study is its emphasis on reproducibility and generalizability. Many previously published lncRNA prognostic models ^34–36^ have been derived from relatively small cohorts, frequently using only the TCGA-LAML dataset, and often lack external validation or systematic robustness analyses. In contrast, our study incorporated two bone marrow and three peripheral blood datasets from three independent cohorts representing clinically distinct AML populations, including adult patients from Beat AML and TCGA-LAML, and pediatric patients from TARGET-AML. Candidate prognostic lncRNAs as well as lncSigAML signature were required to demonstrate reproducible prognostic associations across all datasets, thereby reducing the likelihood of cohort-specific overfitting. Furthermore, sensitivity analyses demonstrated that the prognostic signal was preserved despite substantial differences in patient age, follow-up duration, and disease characteristics between the two cohorts. Together, these findings support the potential generalizability of the proposed lncSigAML signature.

Another distinguishing feature of our study is the interpretability of the selected component lncRNAs and the systematic comparison of multiple survival modeling strategies. First, our multi-step prognostic screening allows the preservation of interpretable properties of the candidate lncRNAs, such as high variability across samples and individual association with survival. Second, we evaluated multiple modeling approaches, including penalized Cox regression, gradient boosting, XGBoost, and random forest under an identical feature set and validation framework. Among these approaches, random survival forests achieved the highest overall predictive performance while demonstrating consistent performance across cross-validation folds. Tree-based ensemble survival models are capable of modeling nonlinear associations and high-order interactions without requiring explicit specification of these relationships, making them well-suited for biologically heterogeneous diseases such as AML. Importantly, rather than assuming that more sophisticated machine-learning methods would outperform conventional approaches, our study provides an empirical comparison that supports the selection of random survival forests for this particular prognostic task.

The comprehensive validation strategy further strengthens the translational relevance of this work. We divided our validation process into mutual-tissue external validation and cross-tissue external validation. We finally performed a meta-analysis to extract the final in addition to conventional validation, we conducted cross-platform validation and robustness to substantial levels of missing expression data. Of importance, model calibration analyses were reported across all validation processes. Calibration is frequently overlooked in prognostic biomarker studies despite being essential for clinical decision-making, as accurate risk estimation requires not only discrimination but also agreement between predicted and observed outcomes. Likewise, sensitivity analyses demonstrated that predictive performance remained relatively stable following imputation of substantial proportions of missing lncRNA measurements, suggesting that the model may retain utility under imperfect real-world data acquisition conditions. Although prospective validation will ultimately be required before clinical implementation, these analyses provide evidence that the proposed framework is resilient across multiple clinically relevant scenarios.

From a clinical perspective, improved molecular risk stratification remains an important objective in AML management ^1^. Current prognostic classification systems rely primarily on cytogenetic abnormalities and recurrent gene mutations but continue to leave many patients within intermediate-risk categories where treatment decisions remain challenging ^3^. Our findings suggest that bone marrow-derived lncRNA expression profiles may provide complementary prognostic information that could potentially augment existing risk stratification systems, as the lncSigAML signature was demonstrated as an independent prognostic factor to clinical variables as well as genetic and cytogenetic subtypes via multivariable Cox analyses and the PLR test. Furthermore, because the signature demonstrated prognostic utility across both pediatric and adult AML cohorts, it may offer broader applicability than models developed exclusively within a single age group. The development of a publicly accessible Shiny application further lowers barriers for independent validation and future clinical investigation by allowing researchers and clinicians to evaluate patient risk without requiring specialized computational expertise.

Several limitations should be acknowledged. First, this study was based on retrospective analyses of publicly available transcriptomic datasets, and prospective validation in independent clinical cohorts will be necessary before routine clinical application. Second, although the signature demonstrated reproducibility across three large AML cohorts, additional validation using geographically diverse populations and different sequencing platforms would further strengthen its generalizability. Third, while the identified lncRNAs exhibited reproducible prognostic associations, the biological functions of several signature components remain incompletely understood and require in vitro functional experiments. Functional studies examining their roles in leukemogenesis, disease progression, and therapeutic resistance will provide important mechanistic insight. Finally, although our multivariable analyses demonstrated that the lncRNA signature provides prognostic information beyond basic clinical variables, future studies should evaluate its incremental value alongside comprehensive molecular risk factors, including ELN risk classification ^1^, recurrent somatic mutations ^37,38^, and measurable residual disease status ^39^.

In conclusion, we developed a bone marrow-derived ten-lncRNA prognostic framework for AML using three cohorts encompassing both pediatric and adult patients. Through systematic biomarker discovery, comprehensive model benchmarking, rigorous validation, and public deployment, this study extends beyond the development of a conventional prognostic signature and proposes a reproducible framework for lncRNA-based prognostic modeling in AML. Future prospective validation and biological characterization of the identified lncRNAs may further support their integration into precision medicine strategies for patients with AML.

## Data Availability

All data produced are available online at https://portal.gdc.cancer.gov/ and https://github.com/biodev/beataml2.0_data.

## TABLE LEGENDS

**Table 1. Multivariable Cox regression analysis of lncSigAML adjusted for clinical variables in the BM1 cohort.** The analysis evaluates the association between lncSigAML risk and overall survival after adjustment for age, sex, and race in the BM1 cohorts. Hazard ratios (HRs), 95% confidence intervals (CIs), and P values are shown.

**Table 2. Multivariable Cox regression analysis of lncSigAML adjusted for genetic subtypes in the BM2 cohort.** The analysis evaluates the independent association of lncSigAML risk with overall survival after adjustment for AML genetic subtypes. Hazard ratios (HRs), 95% confidence intervals (CIs), and P values are shown.

**Table 3. Multivariable Cox regression analysis of lncSigAML adjusted for cytogenetic risk in the PB3 cohort.** The analysis evaluates the independent association of lncSigAML risk with overall survival after adjustment for cytogenetic risk. Hazard ratios (HRs), 95% confidence intervals (CIs), and *P* values are shown.

