## Supplemental data for "lncSigAML: An AI-derived Long Non-coding RNA Prognostic Signature with Multi-Cohort Validation across 2,681 with Acute Myeloid Leukemia Patients"

### SUPPLEMENTAL METHODOLOGY

#### Data preprocessing

*TARGET-AML cohort (BM1 and PB1)*

A total of 3,227 samples from 2,189 unique patients (submitter IDs) were available in the RNA-sequencing dataset. Sample types were identified according to the TCGA sample-type code classification. The distribution of tissue codes is provided in the table below, and detailed definitions of the TCGA sample-type codes are available from the National Cancer Institute Genomic Data Commons.

| Sample code | 03A | 04A | 09A | 10A | 14A | 40A | 41A | 42A | 50A | 85 | Unknown |
|---|---|---|---|---|---|---|---|---|---|---|---|
| No. | 355 | 412 | 1,891 | 31 | 412 | 31 | 25 | 1 | 18 | 50 | 2 |

For the bone marrow (BM1) cohort, samples with tissue code 09A, corresponding to primary tumor tissue from bone marrow, were selected, yielding 1,891 samples from the initial dataset. After removing duplicate samples from the same submitter ID, 1,747 unique samples remained. These samples were subsequently matched to the 2,189 patients in the overall cohort based on submitter ID, resulting in 1,613 patients with corresponding primary tumor bone marrow samples. After restricting to patients with available overall survival data, the final BM1 cohort comprised 1,594 patients.

For the peripheral blood (PB1) cohort, samples with tissue code 03A, corresponding to primary tumor tissue from peripheral blood, were selected, yielding 355 samples. After deduplication by submitter ID, 342 unique samples remained. Matching these samples to the 2,189 patients in the overall cohort based on submitter ID resulted in 314 patients with corresponding primary tumor peripheral blood samples. After restricting to patients with available overall survival data, the final PB1 cohort comprised 312 patients.

Thus, the final BM1 and PB1 cohorts included 1,594 and 312 unique patients, respectively, each with one corresponding primary tumor bone marrow or peripheral blood sample and available overall survival data.

*Beat AML cohorts (BM2 and PB2)*

From the downloaded dataset, a total of 942 patients had collected specimens, including 527 bone marrow, 22 leukapheresis, and 393 peripheral blood samples. These specimens were not restricted to those submitted for RNA sequencing. Among these patients, 707 samples were submitted for RNA sequencing.

For the bone marrow (BM2) dataset, the 527 patients with bone marrow samples were initially selected. These patients were then matched to the 707 patients with RNA-seq data based on submitter ID. As a result, 362 patients remained with both bone marrow specimens and corresponding RNA-seq data. After further restricting to patients with available overall survival data, the final BM2 cohort comprised 356 patients.

For the peripheral blood (PB2) dataset, the 393 patients with peripheral blood samples were initially selected. These patients were subsequently matched to the 707 patients with RNA-seq data based on submitter ID. After restricting the RNA-seq samples to peripheral blood samples, 293 patients remained with both peripheral blood specimens and corresponding RNA-seq data. After restricting to patients with available overall survival data, the final PB2 cohort comprised 277 patients.

Thus, the final BM2 and PB2 cohorts included 356 and 277 unique patients, respectively, each with one corresponding primary tumor bone marrow or peripheral blood sample and available overall survival data.

*TCGA-LAML cohort (PB3)*

The TCGA-LAML cohort comprised patients with peripheral blood samples. From the downloaded data, a total of 151 peripheral blood samples were available for RNA sequencing, while clinical information was available for 200 patients. Of these, 188 patients had available overall survival data. Matching submitter IDs between the 188 patients with available OS data and the 152 RNA-seq samples identified 142 patients with both OS and RNA-seq data. These 142 patients constituted the final TCGA-LAML peripheral blood (PB3) cohort.

**Selecting lncSigAML threshold for detecting low- and high-risk groups**

Because the different cohorts exhibited distinct survival characteristics and distributions of lncSigAML scores, a single predefined cutoff was not applied across all datasets. Instead, cohort-specific thresholds were determined using maximally selected rank statistics (MSRS) implemented through the surv_cutpoint function in the survminer R package. This approach identifies the lncSigAML score threshold that provides the strongest separation in overall survival between the resulting risk groups.

For patient j in cohort i, let $L_{ij}$ denote the lncSigAML score and $T_{ij}$ and $\delta_{ij}$ denote the observed survival time and event indicator, respectively. For a candidate cutoff c, patients are divided into two groups according to $L_{ij} \leq c$ and $L_{ij} > c$. The log-rank statistic compares the observed and expected numbers of events between these two groups at each observed event time.

At event time $t_k$, let $n_{1k}$ and $n_{2k}$ denote the numbers of patients at risk in groups 1 and 2, respectively, and let $d_{1k}$ and $d_{2k}$ denote the corresponding numbers of observed events. The total number at risk and total number of events are $n_k = n_{1k} + n_{2k}$ and $d_k = d_{1k} + d_{2k}$. Under the null hypothesis of identical survival distributions, the expected number of events in group 1 at $t_k$ is

$$E_{1k} = d_k \times (n_{1k} / n_k)$$

The total observed and expected numbers of events in group 1 are

$$O_1 = \Sigma_k \, d_{1k}$$

$$E_1 = \Sigma_k \, E_{1k}$$

The corresponding variance is

$$V_1 = \Sigma_k \, [\, n_{1k} \, n_{2k} \, d_k \, (n_k - d_k) / \{n_k^2 \, (n_k - 1)\} \,]$$

The standardized log-rank statistic for candidate cutoff c is therefore

$$Z(c) = (O_1 - E_1) / \sqrt{V_1}$$

For a two-group comparison, the corresponding test statistic can equivalently be expressed as

$$\chi^2(c) = Z(c)^2$$

The MSRS procedure evaluates the log-rank statistic over candidate lncSigAML cutoffs. The cohort-specific optimal cutoff $c_i$ was defined as the value that maximized the absolute standardized log-rank statistic:

$c_i$ = arg max_c $|Z_i(c)|$

Patients were subsequently classified into low- and high-risk groups according to the cohort-specific cutoff:

$R_{ij} = 0$ if $L_{ij} \leq c_i$ (low risk); $R_{ij} = 1$ if $L_{ij} > c_i$ (high risk)

The resulting binary risk-group variable $R_{ij}$ was used for subsequent survival and differential expression analyses within each cohort. The use of cohort-specific MSRS thresholds was motivated by differences in survival characteristics and lncSigAML score distributions across cohorts. In addition, the prognostic association of lncSigAML may not be uniform across the full range of scores. A data-driven threshold therefore provides a consistent statistical procedure for identifying the score value that best separates patients according to overall survival within each independent cohort.

Because multiple candidate cutoffs are evaluated during the MSRS procedure, the significance of the maximally selected statistic was interpreted within the MSRS framework rather than as an unadjusted log-rank test at a prespecified cutoff.

**Sensitivity Analyses**

First, a separate random survival forest model was trained using the Beat AML cohort (BM2) and applied to the TARGET-AML cohort (BM1). Model discrimination power was assessed using the C-index, and 1-, 3-, and 5-year calibrations were evaluated using the calibration framework described above. In the reverse validation direction, a model trained on Beat AML BM2 (n = 356) was applied to the TARGET-AML BM1 cohort (n = 1,594) as the independent test set

**Performance under missingness analysis**

A missing-data sensitivity analysis was performed to evaluate stability of lncSigAML to incomplete expression measurements. We conducted this stress test experiment on the BM2 dataset. Missingness was then introduced randomly into the lncRNA expression features of the held-out test set. In the current analysis, missingness was introduced at four missingness thresholds: 10%, 20%, 30%, 40%, and 50% in the test set. Missing values were imputed using multivariate imputation by chained equations (MICE) [1-4] with predictive mean matching, implemented using the mice package. One imputed dataset was generated using five imputation iterations. After imputation, the trained random survival forest model was applied to the imputed test data. Model discrimination was evaluated using the C-index, and 1-, 3-, and 5-year calibrations was evaluated using the calibration framework described above.

**Meta-analysis of hazard ratio**

Because the four independent test cohorts differed in demographic characteristics, clinical features, and RNA-sequencing technologies, cohort-specific estimates may vary despite reflecting a common underlying prognostic association. To assess the consistency of the prognostic effect of lncSigAML across cohorts, we performed meta-analysis of the hazard ratio (HR), survival probability, and log2 fold-change estimates. Dataset-specific results were also retained and reported separately to allow assessment of cohort-level heterogeneity and consistency.

For the HR meta-analysis, hazard ratios were pooled using an inverse-variance weighting approach. Because the sampling distribution of the HR is asymmetric, the analysis was performed on the natural logarithm of the HR. For each cohort (i), the estimated log-hazard ratio was calculated as

$$\theta_i = \log(HR_i)$$

with its corresponding standard error ((SE_i)) obtained from the Cox proportional hazards model. The variance of each estimate was calculated as

$$V_i = SE_i^2$$

Under a fixed-effect model, each cohort was assigned an inverse-variance weight,

$$w_i = \frac{1}{V_i} = \frac{1}{SE_i^2}$$

such that cohorts with more precise HR estimates contributed greater weight to the pooled effect. The pooled log-hazard ratio was then calculated as

$$\hat{\theta} = \frac{\sum_{i=1}^{k} w_i \theta_i}{\sum_{i=1}^{k} w_i}$$

where (k) is the number of independent cohorts. The corresponding pooled hazard ratio was obtained by exponentiating the pooled log-hazard ratio:

$$HR_{pooled} = \exp(\hat{\theta})$$

**Meta-analysis of survival probabilities**

To evaluate the consistency of the survival differences between lncSigAML-defined low- and high-risk groups across the four independent test cohorts, survival probabilities were meta-analyzed at prespecified time points. Because survival probabilities are bounded between 0 and 1, the analysis was performed on the logit-transformed survival probability to improve the normality and stability of the estimates.

For each cohort (i) and risk group (g), the estimated survival probability at a given time point (t), denoted $S_{i,g}(t)$, was obtained from the Kaplan–Meier survival curve. The survival probability was transformed to the logit scale as

$$log\left(\frac{S_{i,g}(t)}{1 - S_{i,g}(t)}\right)$$

The corresponding standard error was obtained from the Kaplan–Meier estimate using Greenwood's variance estimator and propagated to the logit scale using the delta method. Specifically,

$$SE_{\hat{\theta}_{i,g}(t)} \approx \frac{SE_{S_{i,g}(t)}}{S_{i,g}(t)[1 - S_{i,g}(t)]}$$

The variance of each transformed estimate was then calculated as

$$V_{i,g}(t) = SE_{\theta_{i,g}(t)}{}^{2}$$

and inverse-variance weights were assigned as

$$w_{i,g}(t) = \frac{1}{V_{i,g}(t)}$$

The pooled logit-transformed survival probability was calculated as

$$\frac{exp\hat{\theta}_g(t)}{1 + exp\hat{\theta}_g(t)}$$

The standard error of the pooled estimate was calculated as

$$\sqrt{\frac{1}{\sum_{i=1}^{k} w_{i,g}\,(t)}},$$

and the 95% confidence interval was calculated on the logit scale as

$$\hat{\theta}_g(t) \pm 1.96 \times SE\hat{\theta}_g(t).$$

with the resulting confidence limits transformed back to the probability scale using the inverse-logit transformation.

This procedure was performed separately for the low- and high-risk groups at each prespecified time point. The pooled survival probabilities therefore represent the consensus survival estimates across the independent test cohorts while accounting for the precision of the cohort-specific estimates. Dataset-specific Kaplan–Meier estimates were retained and reported separately to allow direct assessment of between-cohort consistency.

**Partial Likelihood Ratio (PLR) Tests**

To determine whether lncSigAML provided incremental prognostic information beyond the variables included in the current AML prognostication model, we compared nested Cox proportional hazards regression models using a partial likelihood ratio (PLR) test. The reduced model (M2) included the established clinical and/or molecular prognostic variables in the current prognostication model, whereas the full model (M1) additionally included lncSigAML:

M1: OS ~ current prognostic variables+lncSigAML

M2: OS ~ Age+ELN risk group+WBC count

where overall survival (OS) was modeled using survival time and censoring status.

The two models were fitted using the Cox proportional hazards model, and their maximized partial log-likelihoods were compared. The PLR statistic was calculated as

$$LR = 2(\ell_{M1} - \ell_{M2})$$

where $\ell_{M1}$ and $\ell_{M2}$ denote the maximized partial log-likelihoods of the full and reduced models, respectively. Under the null hypothesis that lncSigAML does not provide additional prognostic information after accounting for the variables in the current model, the LR statistic approximately follows a chi-square distribution with degrees of freedom equal to the number of additional parameters introduced by lncSigAML. Because lncSigAML was modeled as a single covariate, the comparison had one degree of freedom.

A significant PLR test was interpreted as evidence that inclusion of lncSigAML significantly improved the prognostic model beyond the established prognostic variables. The PLR test therefore evaluated the incremental prognostic value of lncSigAML rather than its association with OS considered independently. The likelihood ratio statistic, degrees of freedom, and corresponding two-sided P value were reported for each model comparison.

**Differential expression analyses (DEA)**

To characterize molecular differences between the lncSigAML-defined low- and high-risk groups, differential gene expression analysis was performed independently within each test cohort. The analysis was restricted to the independent test cohorts and did not use the training cohort, thereby avoiding the incorporation of training data into downstream molecular characterization and preserving the independence of the validation analyses. Differential expression was assessed using DESeq2, an R/Bioconductor package designed for the analysis of count-based RNA-sequencing data. DESeq2 models gene-level read counts using a negative binomial distribution and accounts for differences in sequencing depth and biological variability between samples through size-factor normalization and dispersion estimation. Compared with analyses based directly on normalized expression values, this framework appropriately models the mean–variance relationship inherent to RNA-sequencing count data and provides statistical inference for differential expression.

Within each test cohort, patients were stratified into low- and high-risk groups according to the cohort-specific lncSigAML cutoff described above. For each gene $g$, the observed read count for sample $j$, $K_{gj}$, was modeled using a negative binomial generalized linear model:

$$K_{gj} \sim NB(\mu_{gj}, \alpha_g),$$

Where $\mu_{gj}$ is the expected normalized count and $\alpha_g$ is the gene-specific dispersion parameter. The expected count was modeled as

$$log(\mu_{gj}) = log(s_j) + \beta_{0g} + \beta_{1g} R_j,$$

Where $s_j$ denotes the sample-specific size factor, $\beta_{0g}$ represents the baseline expression level of gene $g$, $R_j$ is the risk-group indicator, and $\beta_{1g}$ represents the expression difference between the high- and low-risk groups on the natural-log scale. Specifically, $R_j = 0$ for the low-risk group and $R_j = 1$ for the high-risk group. Thus,

$$\beta_{1g} = log\left(\frac{\mu_{g,high}}{\mu_{g,low}}\right),$$

and the corresponding log2 fold change reported by DESeq2 is

$$log_2(FC_g) = \frac{\beta_{1g}}{\ln(2)}.$$

Differential expression analysis was performed separately for each test cohort rather than combining the RNA-sequencing data across cohorts. This cohort-specific approach accounts for differences in patient characteristics, sample composition, sequencing platforms, and other cohort-specific technical or biological factors. Within each cohort, *P* values were adjusted for multiple testing across all analyzed genes using the Benjamini–Hochberg procedure, with a false discovery rate (FDR)-adjusted *P* value used to assess statistical significance. The resulting cohort-specific effect estimates were subsequently used as inputs for the meta-analysis to identify genes showing consistent differential expression across independent datasets.

**Meta-analysis of DEA results**

To identify genes showing consistent differential expression across independent test cohorts, we performed a meta-analysis of gene-level differential expression results generated using DESeq2. Because the cohorts differed in demographic characteristics, clinical features, and sequencing technologies, cohort-specific effect estimates were retained while a meta-analytic estimate was calculated to assess the overall direction and magnitude of differential expression.

For each gene $j$in cohort $i$, the estimated effect size was defined as the DESeq2 log2 fold change:

$$\theta_{ij} = \log_2(FC_{ij}),$$

where positive values indicate higher expression in the high-risk group relative to the low-risk group, and negative values indicate lower expression. The corresponding standard error, $SE_{ij}$, was obtained from the DESeq2 model. The variance of each estimate was calculated as

$$v_{ij} = SE_{ij}^2.$$

Gene-level effect estimates were then combined across cohorts using inverse-variance weighting. For each gene, the weight assigned to cohort $i$was

$$w_{ij} = \frac{1}{v_{ij}} = \frac{1}{SE_{ij}^2}.$$

The pooled log2 fold change for gene $j$was calculated as

$$\hat{\theta}_j = \frac{\sum_{i=1}^{k} w_{ij}\,\theta_{ij}}{\sum_{i=1}^{k} w_{ij}},$$

where $k$represents the number of cohorts with an evaluable estimate for gene $j$. The standard error of the pooled estimate was calculated as

$$SE(\hat{\theta}_j) = \sqrt{\frac{1}{\sum_{i=1}^{k} w_{ij}}}.$$

The corresponding 95% confidence interval was calculated as

$$\hat{\theta}_j \pm 1.96 \times SE(\hat{\theta}_j),$$

and the corresponding meta-analytic $Z$-statistic was calculated as

$$Z_j = \frac{\hat{\theta}_j}{SE(\hat{\theta}_j)}.$$

Two-sided $P$-values were obtained from the standard normal distribution based on the corresponding $Z$-statistics. Multiple-testing correction was performed across genes using the Benjamini–Hochberg procedure to obtain false discovery rate (FDR)-adjusted $P$-values.

The resulting pooled log2 fold change represents the overall direction and magnitude of differential expression across the independent cohorts, with greater weight assigned to cohorts providing more precise estimates. Positive pooled log2 fold changes indicate genes consistently upregulated in the high-risk group, whereas negative values indicate genes consistently downregulated in the high-risk group. Cohort-specific log2 fold changes and statistical results were retained and reported alongside the meta-analysis to assess the consistency of individual gene-level effects across datasets.

**Construction of the model-ready lncRNA feature matrix**

For each patient, normalized expression values of the eight selected lncRNAs were extracted from the BeatAML and TARGET-AML expression matrices. Cohort-specific model-ready datasets were generated by combining sample identifiers, OS time, OS status, and the eight lncRNA expression features. The BeatAML and TARGET-AML model-ready datasets were harmonized to retain shared columns and were then concatenated for combined-cohort modeling. Before machine-learning analysis, the lncRNA features were standardized at the patient level. Specifically, for each patient, the mean expression across the ten lncRNAs was subtracted from each lncRNA expression value and divided by the standard deviation across the ten lncRNAs. This generated a standardized ten -feature lncRNA expression profile for each patient. Patients with incomplete OS data or missing values for any of the ten lncRNA features were removed before model training and validation.

**Machine-learning model selection by cross-validation**

Multiple survival prediction models were evaluated using the same standardized ten -lncRNA feature matrix. Candidate models included Cox proportional hazards regression LASSO-Cox regression, Elastic-net Cox regression, Random survival forest, XGBoost Cox survival model, and Gradient boosting Cox model. Model comparison was performed using five-fold cross-validation in the combined BeatAML and TARGET-AML cohort. Cross-validation folds were generated using stratification by OS event status. For each fold, candidate models were trained on four-fifths of the cohort and evaluated on the held-out test fold. Model discrimination was assessed using Harrell's concordance index. The mean and standard deviation of the C-index across the five folds were calculated for each candidate model. The model with the highest cross-validated C-index was selected for downstream prognostic modeling, calibration, risk stratification, cross-dataset validation, and sensitivity analyses (Table 2). Model comparison was performed using five-fold cross-validation. Cross-validation folds were generated using stratification by OS event status. For each fold, candidate models were trained on four-fifths of the cohort and evaluated on the held-out test fold. Model discrimination was assessed using Harrell's concordance index (C-index) [5]. The mean and standard deviation of the C-index across the five folds were calculated for each candidate model. The model with the highest cross-validated C-index was selected for downstream prognostic modeling, risk stratification, external, and cross-tissue validation.

For each fold, candidate models were trained on four-fifths of the cohort and evaluated on the held-out test fold. Model discrimination was assessed using Harrell's concordance index (C-index) [5]. The mean and standard deviation of the C-index across the five folds were calculated for each candidate model. The model with the highest cross-validated C-index was selected for downstream prognostic modeling, risk stratification, external, and cross-tissue validation.

demonstrates the overall workflow to demonstrate the prognostic effect of lncSigAML. Patients in the BM1 (N = 1,594) were stratified into low-risk (n = 1,036) and high-risk (n = 558) groups based on the maximally selected rank statistics. Kaplan-Meier survival analysis confirmed a statistically significant difference in OS between the two risk groups (log-rank $P < 0.001$)**,** wherein high-risk patients exhibited markedly worse outcomes compared with their low-risk counterparts, corresponding to a hazard ratio

**Candidate model specifications**

The Cox proportional hazards model was fit using the ten lncRNA predictors as continuous covariates. LASSO-Cox and elastic-net Cox models were implemented using glmnet. For LASSO-Cox regression, the mixing parameter was set to alpha = 1. For elastic-net Cox regression, the mixing parameter was set to alpha = 0.5. The regularization parameter was selected using internal cross-validation within the training set, and predictions were generated using lambda.min. Random survival forest models were implemented using the randomForestSRC package. The survival outcome was specified as: $Surv(OS\ time, OS\ status)$. The ten standardized lncRNA expression features were used as predictors. The number of trees was set to 1,000. XGBoost survival models were implemented using the Cox survival objective. The model was trained with 200 boosting rounds, maximum tree depth of 3, learning rate of 0.05, subsampling fraction of 0.8, and column subsampling fraction of 0.8. Gradient boosting models were implemented using the gbm package with Cox proportional hazards loss. The number of trees was set to 500 and interaction depth was set to 3.

**Internal validation and calibration**

Internal validation of the final random survival forest model was performed using five-fold cross-validation in the combined cohort. For each fold, the model was trained on the training subset and applied to the held-out test subset. Out-of-fold predictions were retained for all patients.

Discrimination was evaluated using Harrell's C-index. Calibration of predicted 1-year survival probability was assessed using pooled out-of-fold predictions. A Cox proportional hazards calibration model was fit using the random survival forest-derived risk score as the predictor. Calibration curves were generated using the cph and calibrate functions from the rms package.

Observed survival probabilities were estimated using Kaplan–Meier-based calibration. Bootstrap resampling with B = 40 resamples was used to estimate bias-corrected calibration. Calibration curves plotted predicted 1-year survival probability against observed 1-year survival probability, with the 45-degree line representing perfect calibration.

**Missing-data sensitivity analysis**

A missing-data sensitivity analysis was performed to evaluate robustness of the eight-lncRNA random survival forest model to incomplete expression measurements. Five-fold cross-validation was repeated in the combined cohort. Within each fold, the model was trained using complete training data. Missingness was then introduced randomly into the eight lncRNA expression features of the held-out test set. In the current analysis, missingness was introduced at a rate of 80% for each selected lncRNA feature in the test set. Missing values were imputed using multivariate imputation by chained equations with predictive mean matching, implemented using the mice package. One imputed dataset was generated using five imputation iterations. After imputation, the trained random survival forest model was applied to the imputed test data. Model discrimination was evaluated using the C-index, and 1-year calibration was assessed using the same calibration workflow described above.

**Risk stratification and survival analysis**

Patients were stratified into high-risk and low-risk groups based on the median random survival forest-derived risk score. Patients with risk scores above the median were assigned to the high-risk group, whereas patients with risk scores less than or equal to the median were assigned to the low-risk group. Kaplan–Meier survival curves were generated to compare OS between high-risk and low-risk patients. Survival differences were evaluated using the log-rank test. Hazard ratios and 95% confidence intervals were estimated using Cox proportional hazards regression with risk group as the predictor. For cross-dataset validation analyses, risk groups were defined using model-derived risk scores in the corresponding test cohort.

**Exploratory Survival Analysis**

Kaplan–Meier survival analysis was conducted for selected lncRNAs. Samples were stratified into high- and low-expression groups based on median expression levels. Survival differences were evaluated using the log-rank test.

**Functional analyses**

Differential expression, GSEA, GO enrichment, KEGG, Hallmark pathways, Leukemia stem cell signatures, Immune infiltration

**Web application**

The prognostic model was deployed using R Shiny, Docker, Python backend. Input: normalized TPM matrix. Output: Risk score, Risk group, Survival curves, Nomogram, Forest plot, Downloadable report.

**Sensitivity Analyses**

***Cross-platform validation experiment***

To evaluate generalizability across independent AML cohorts, bidirectional cross-platform validation of the 2 bone marrow datasets (BM1-2) was performed. First, a random survival forest model was trained using the TARGET-AML cohort (BM1) and tested in the Beat AML cohort (BM2). Second, a separate random survival forest model was trained using the Beat AML cohort (BM2) and tested in the TARGET-AML cohort (BM1) (**Figure 3A**). For each validation direction, the same ten-lncRNA feature set and modeling framework were used. The trained model was applied to the independent test cohort to generate patient-level risk scores. Model discrimination was assessed using the C-index, and 3-year calibration was evaluated using the calibration framework described above.

**SUPPLEMENTAL FIGURES**

**A** Significant OS-associated lncRNAs

| Gene | HR | P value | C index | Adjusted P | Gene Variance |
|---|---|---|---|---|---|
| HOXA-AS3 | 1.18123025 | 7.46E-17 | 0.595004777 | 1.62E-14 | 2.083436534 |
| LINC01122 | 1.209253016 | 1.49E-15 | 0.589115814 | 3.23E-13 | 1.689322074 |
| HOXA10-AS | 1.149044892 | 1.31E-14 | 0.58845389 | 2.85E-12 | 2.445150302 |
| SOCS2-AS1 | 1.22130253 | 1.08E-14 | 0.587360277 | 2.34E-12 | 1.672899857 |
| LINC01108 | 1.177461026 | 2.41E-15 | 0.581904441 | 5.22E-13 | 1.757162255 |
| HOTAIRM1 | 1.210435095 | 9.27E-11 | 0.581748314 | 2.01E-08 | 1.439228652 |
| HOXA11-AS | 1.148789485 | 2.46E-10 | 0.577205501 | 5.34E-08 | 1.802962313 |
| KCNJ2-AS1 | 1.226936721 | 3.05E-11 | 0.574296634 | 6.62E-09 | 1.290862927 |
| MEIS1-AS2 | 1.163673488 | 2.32E-11 | 0.573999488 | 5.04E-09 | 1.787778193 |
| TSPEAR-AS1 | 1.187511362 | 5.89E-10 | 0.571092059 | 1.28E-07 | 1.35077536 |
| LINC00355 | 1.173024744 | 7.80E-12 | 0.570019311 | 1.69E-09 | 1.455755157 |
| LINC00299 | 1.198608866 | 3.31E-08 | 0.569928656 | 7.18E-06 | 1.340298938 |
| CYP4F26P | 1.118606152 | 2.77E-08 | 0.565111721 | 6.02E-06 | 1.947276245 |
| HOXA-AS2 | 1.194922034 | 2.22E-09 | 0.565094454 | 4.81E-07 | 1.301308878 |
| ST3GAL6-AS1 | 0.848766347 | 4.24E-08 | 0.564996604 | 9.19E-06 | 1.430411913 |
| LINC01013 | 1.170967769 | 1.09E-07 | 0.564081422 | 2.37E-05 | 1.31290007 |
| MIR17HG | 1.18653205 | 1.59E-06 | 0.56347346 | 3.46E-04 | 1.315772439 |
| FOXD2-AS1 | 1.184989766 | 1.23E-07 | 0.562643177 | 2.67E-05 | 1.369135767 |
| FLG-AS1 | 1.169512398 | 1.56E-08 | 0.560280397 | 3.39E-06 | 1.481316246 |
| PROSER2-AS1 | 0.909966085 | 2.07E-06 | 0.560052321 | 4.49E-04 | 2.205579716 |
| MEF2C-AS1 | 1.190699649 | 2.77E-07 | 0.559310534 | 6.00E-05 | 1.325657792 |
| LINC00477 | 1.121547747 | 3.71E-08 | 0.558856541 | 8.04E-06 | 1.800749861 |
| ADARB2-AS1 | 0.844043228 | 2.23E-07 | 0.557246339 | 4.83E-05 | 1.517487906 |
| AP001042.1 | 1.155100936 | 7.83E-07 | 0.550288944 | 0.000169878 | 1.386549762 |

Genomic organization of the 10-lncRNA signature

**B** *LINC01122*

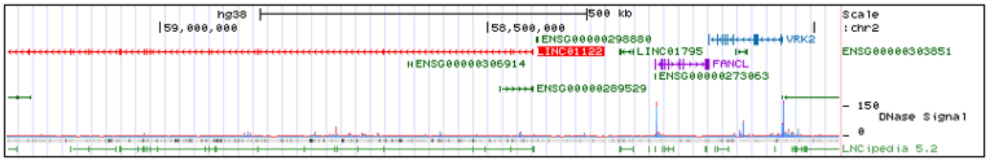


**C** *SOCS2-AS1*

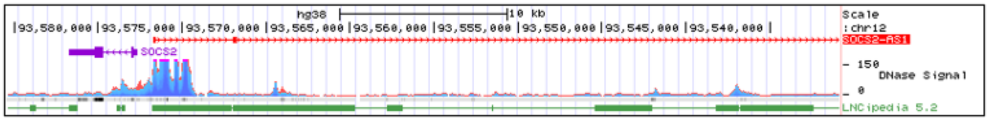


**D** *LINC01108*

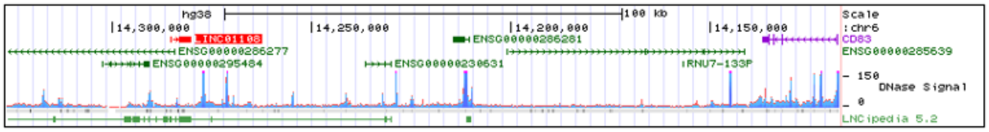

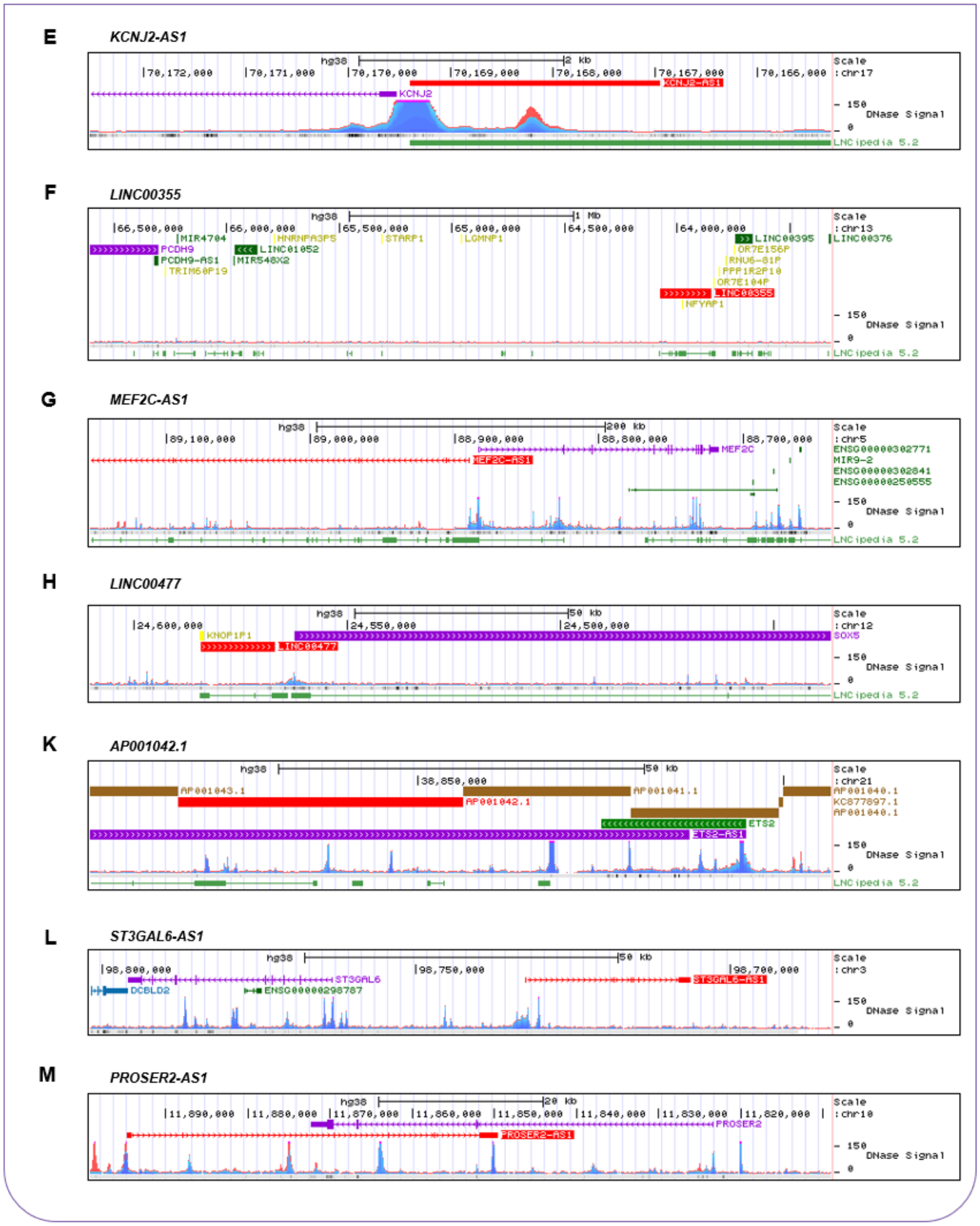


**Figure S1. The 24 OS-associated lncRNAs in the BM1 cohort, Related to Figure 2**

(A) The 24 OS-associated lncRNAs identified from the most highly variable lncRNAs in the BM1 cohort using univariate Cox proportional hazards regression with Bonferroni correction ($P < 0.001$).

(B-H) Gene track view of the genomic region containing the human gene loci of top ten OS-associated lncRNAs. Shown are GENCODE V50 track of coding and non-coding genes and the ENCODE DNase I Hypersensitivity Signal from multiple cell lines.

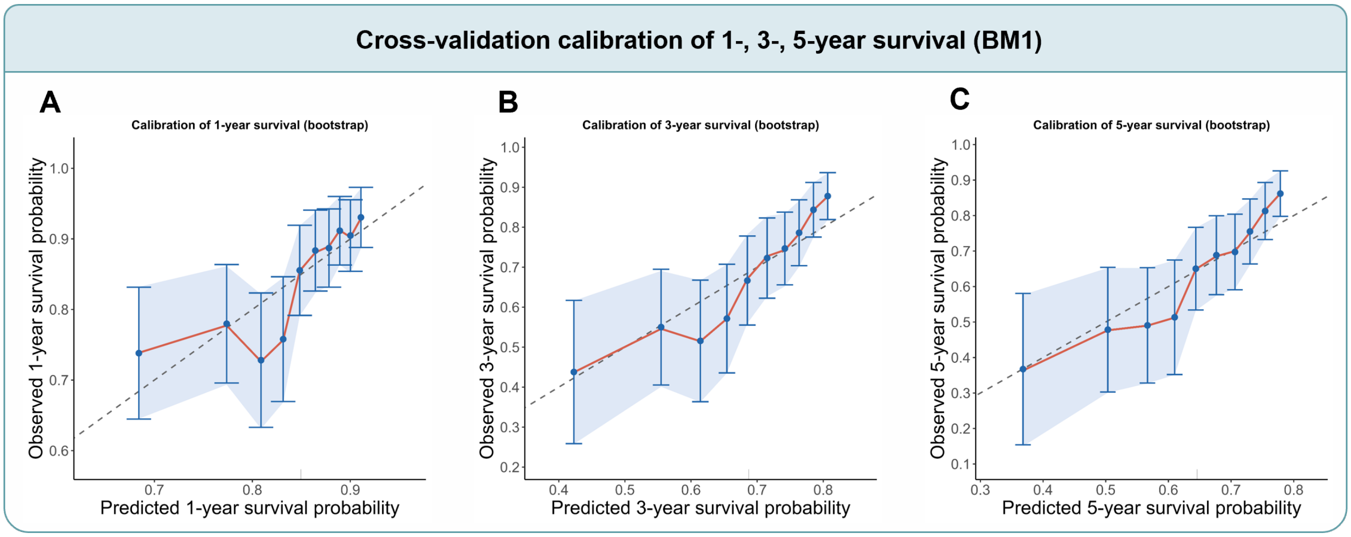


**Figure S2. Cross-validation calibration of lncSigAML in the BM1 cohort, Related to Figure 3**

(A–C) Calibration plots of predicted 1-, 3-, and 5-year overall survival probabilities, respectively, based on cross-validated risk predictions in the BM1 cohort. Calibration was assessed using bootstrap resampling. The x-axis represents the predicted survival probability, and the y-axis represents the observed survival probability. The dashed diagonal line indicates perfect calibration, while the calibration curves and corresponding confidence intervals illustrate the agreement between predicted and observed survival probabilities.

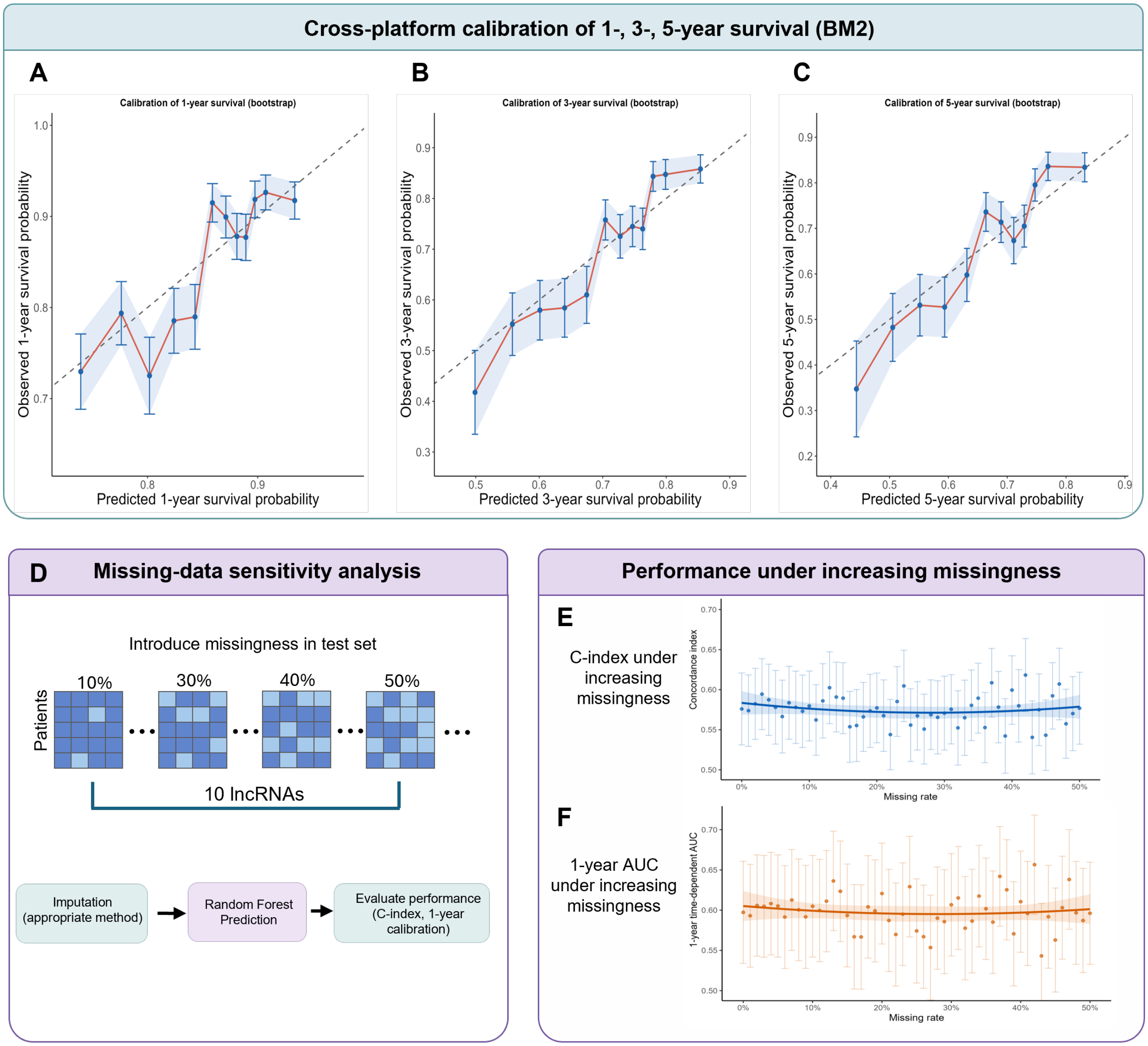
Cross-platform calibration of 1-, 3-, 5-year survival (BM2)
A
Calibration of 1-year survival (bootstrap)
Observed 1-year survival probability
Predicted 1-year survival probability
B
Calibration of 3-year survival (bootstrap)
Observed 3-year survival probability
Predicted 3-year survival probability
C
Calibration of 5-year survival (bootstrap)
Observed 5-year survival probability
Predicted 5-year survival probability
D
Missing-data sensitivity analysis
Introduce missingness in test set
Patients
10%
30%
40%
50%
10 lncRNAs
Imputation (appropriate method)
Random Forest Prediction
Evaluate performance (C-index, 1-year calibration)
Performance under increasing missingness
E
C-index under increasing missingness
Concordance index
Missing rate
F
1-year AUC under increasing missingness
1-year time-dependent AUC
Missing rate

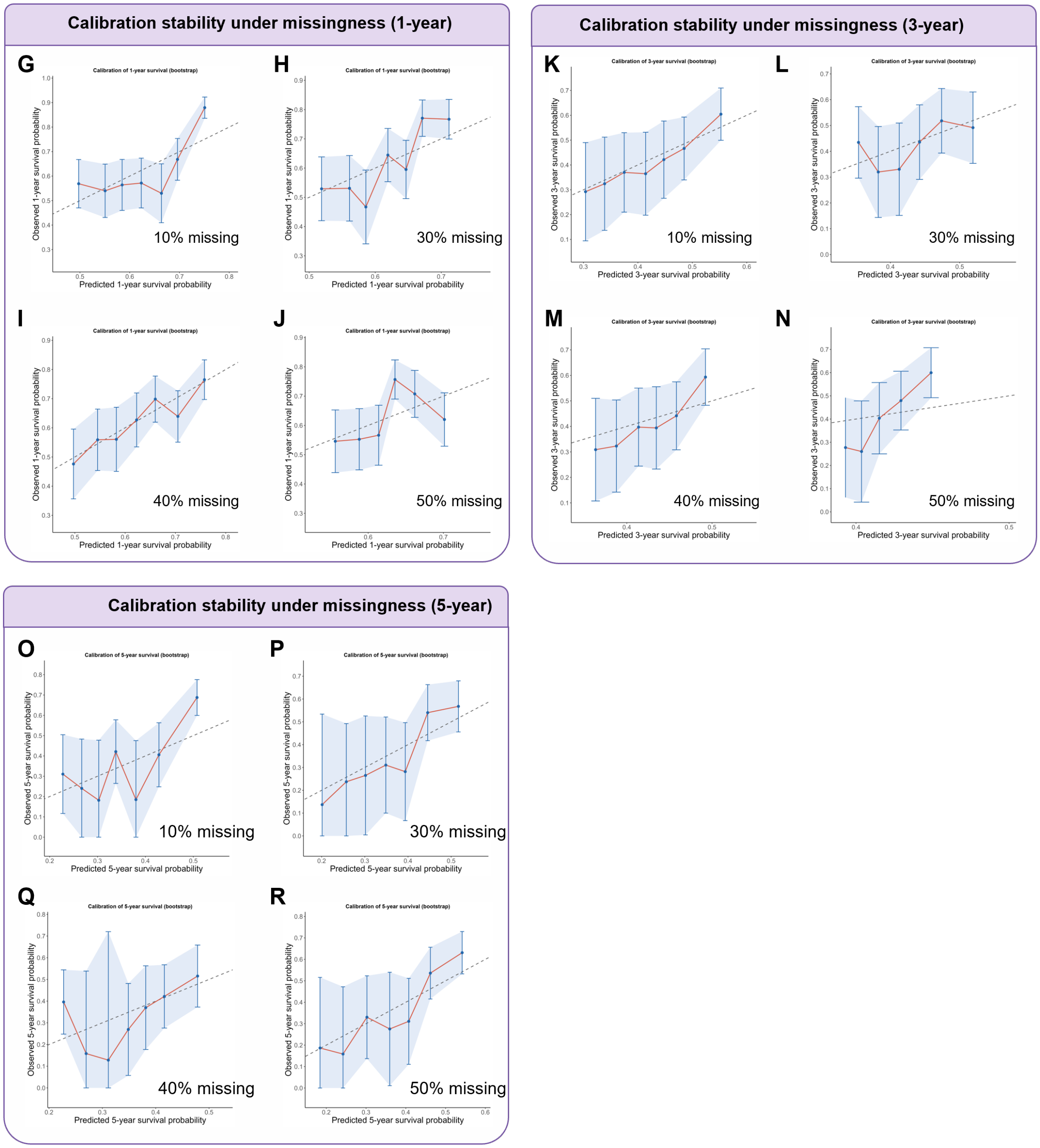


**Figure S3. Cross-platform calibration and missing-data sensitivity analysis of lncSigAML-RFM, Related to Figure 4**

(A-C) Calibration plots for predicted 1-, 3-, and 5-year overall survival probabilities, respectively, generated using bootstrap resampling in the BM2 cohort. The x-axis represents the predicted survival probability, and the y-axis represents the observed survival probability. The dashed diagonal line indicates perfect calibration, while the calibration curves and corresponding confidence intervals show the agreement between predicted and observed survival probabilities.

(D) Schematic of the missing-data perturbation analysis. Increasing levels of missingness (10%, 30%, 40%, and 50%) were introduced into the test set across the 10 lncRNAs comprising the lncSigAML signature. Missing expression values were imputed using the appropriate imputation method, followed by risk prediction using the pretrained Random Forest model. Model performance was subsequently evaluated using the concordance index (C-index) and 1-year calibration.

(E-F) lncSigAML performance under increasing levels of missingness. The C-index (E) and 1-year time-dependent area under the receiver operating characteristic curve (AUC) (F) were evaluated across increasing missing rates. Points represent the estimated performance, and error bars indicate the corresponding 95% confidence intervals.

(G–J) Calibration of predicted survival probabilities under increasing levels of missingness at 1 (G-J), 3 (K-N), and 5 (O-R) years, respectively. Calibration plots are shown for 10% (G,K,O), 30% (H,L,P), 40% (I,M,Q), and 50% (J,N,R) missingness. The x-axis represents the predicted survival probability, and the y-axis represents the observed survival probability. The dashed diagonal line indicates perfect calibration, while the calibration curves and confidence intervals illustrate the agreement between predicted and observed survival probabilities across different levels of missingness.

### SUPPLEMENTAL TABLE LEGENDS

**Table S1. Clinical and demographic characteristics of patients across the five AML datasets.** The table summarizes key demographic and clinical characteristics of patients included in the BM1, BM2, PB1, PB2, and PB3 datasets, including cohort size and available clinical, genetic, cytogenetic, and survival characteristics.

**Table S2. Highly variable lncRNAs identified in the BM1 cohort.** The table summarizes the 217 lncRNAs selected based on the standard deviation of log-transformed read counts across BM1 samples, together with their univariate Cox regression results, including hazard ratio (HR), *P* value, concordance index (C-index), Bonferroni-adjusted *P* value, and gene expression variance.

**Table S3. Multivariable Cox regression analysis of lncSigAML adjusted for clinical variables across four independent validation datasets.** The analysis evaluates the association between lncSigAML risk and overall survival after adjustment for age, sex, and race in the BM2, PB1, PB2, and PB3 cohorts. Hazard ratios (HRs), 95% confidence intervals (CIs), and *P* values are shown.

**Table S4. Multivariable Cox regression analysis of lncSigAML adjusted for genetic subtypes in the PB2 cohort.** The analysis evaluates the independent association of lncSigAML risk with overall survival after adjustment for AML genetic subtypes. Hazard ratios (HRs), 95% confidence intervals (CIs), and *P* values are shown.

**Table S5. Differentially expressed genes between lncSigAML risk groups across independent AML cohorts.** The table presents the results of gene-level differential expression analysis between high- and low-risk lncSigAML groups for each independent test dataset (BM2, PB1, PB2, and PB3), together with the meta-analysis of differential expression across the four cohorts. Values include log2 fold change (Log2FC), standard error (SE), Z statistic, *P* value, and false discovery rate (FDR) are shown.

**Table S6. Gene set enrichment analysis (GSEA) of lncSigAML risk groups.** GSEA was performed using the C6 oncogenic gene set collection from the Molecular Signatures Database (MSigDB) to identify pathways enriched in the high- and low-risk lncSigAML groups. The table reports the gene set size (setSize), enrichment score (ES), normalized enrichment score (NES), nominal *P* value, false discovery rate (FDR), nominal significance metric (Q), enrichment rank (Rank), leading-edge subset, and core enrichment genes.

### QUANTITATION AND STATISTICAL ANALYSIS

All analyses were performed in R language version 4.3.1 (The R Foundation, Vienna, Austria). Data preprocessing and visualization were conducted using the *tidyverse*, *dplyr*, *tidyr*, *readr*, and *ggplot2* packages. Survival analyses were performed using the survival and survminer packages. Penalized Cox proportional hazards models were implemented using *glmnet*. Survival random forest models were developed using *randomForestSRC*, XGBoost survival models using *xgboost*, and gradient boosting Cox models using *gbm*. Cross-validation folds were generated using *caret*. Calibration analyses were performed using *rms*. Missing data were imputed using *mice*. All statistical tests were two-sided unless otherwise specified, and a P value < 0.05 was considered statistically significant.

### CONTACT FOR RESOURCE SHARING

2. Buuren Sv. Flexible Imputation of Missing Data, Second Edition

; Published2018.

3. Stef van Buuren KG-O. mice: Multivariate Imputation by Chained Equations in R . *Journal of Statistical Software*. 2011.

4. Van Buuren S, Brand JPL, Groothuis-Oudshoorn CGM, Rubin DB. Fully conditional specification in multivariate imputation. *Journal of Statistical Computation and Simulation*. 2006;76(12):1049-1064.

5. Harrell FE, Jr., Califf RM, Pryor DB, Lee KL, Rosati RA. Evaluating the yield of medical tests. *JAMA*. 1982;247(18):2543-2546.
